# Incorporating epidemiological data into the genomic analysis of partially sampled infectious disease outbreaks

**DOI:** 10.1101/2024.10.31.24316484

**Authors:** Jake Carson, Matt Keeling, Paolo Ribeca, Xavier Didelot

## Abstract

Pathogen genomic data is increasingly being used to investigate transmission dynamics in infectious disease outbreaks. Combining genomic data with epidemiological data should substantially increase our understanding of outbreaks, but this is highly challenging when the outbreak under study is only partially sampled, so that both genomic and epidemiological data are missing for intermediate links in the transmission chains. Here we present a new dynamic programming algorithm to perform this task efficiently. We implement this methodology into the well-established TransPhylo framework to reconstruct partially sampled outbreaks using a combination of genomic and epidemiological data. We use simulated datasets to show that including epidemiological data can improve the accuracy of the inferred transmission links compared to inference based on genomic data only. This also allows us to estimate parameters specific to the epidemiological data (such as transmission rates between particular groups) which would otherwise not be possible. We then apply these methods to two real-world examples. Firstly, we use genomic data from an outbreak of tuberculosis in Argentina, for which data was also available on the HIV status of sampled individuals, in order to investigate the role of HIV co-infection in the spread of this tuberculosis outbreak. Second, we use genomic and geographical data from the 2003 epidemic of avian influenza H7N7 in the Netherlands to reconstruct its spatial epidemiology. In both cases we show that incorporating epidemiological data into the genomic analysis allows us to investigate the role of epidemiological properties in the spread of infectious diseases.

## INTRODUCTION

Over the past decade there has been considerable research interest and methodological development in the analysis of pathogen genomic sequences to reconstruct the transmission events that occurred during an infectious disease outbreak (Jombart et al., 2014; Croucher and Didelot, 2015; Campbell et al., 2018; Duault et al., 2022). Additional epidemiological data about the infected hosts is often available, and it can be useful to integrate such data into the genomic analysis for two complementary reasons. Firstly, it should allow the transmission trees to be reconstructed more precisely when using genomic and epidemiological data compared to using genomic data alone. An example of this was provided by the reconstruction of a tuberculosis outbreak in British Columbia, in which the matrix of who-infected-whom probabilities contained less uncertainty when geographic data and measures of infectiousness (provided by smear and skin tests) were used as additional input (Didelot et al., 2014; Biek et al., 2015; Hatherell et al., 2016). Secondly, using data on epidemiological properties can enable inference on the correlation between transmission and these epidemiological properties. For example, individuals with high-risk behaviours contribute disproportionately to the spread of sexual diseases such as gonorrhoea (Chan et al., 2012; Fingerhuth et al., 2016; Whittles et al., 2019). Integrating behavioural data into a transmission analysis could help quantify this effect, which could be used in predictive models for example to inform the design of control measures such as targeted vaccination (Craig et al., 2015; Whittles et al., 2020, 2022).

Previous attempts have been made to integrate epidemiological and genomic data into outbreak reconstructions. The simplest case occurs if we assume that all cases of the outbreak have been sampled and are therefore present in the transmission tree. In this case the likelihood of the genomic data can simply be multiplied by the likelihood of the epidemiological data (Ypma et al., 2012; Morelli et al., 2012; Didelot et al., 2014; Hall et al., 2015). The epidemiological component of the likelihood is easy to compute as a product over all links in the transmission tree (Ypma et al., 2012). Each link represents an infection from a sampled infector to a sampled infectee, and as epidemiological data are available for both hosts, the contribution to the likelihood is analytically tractable. However, the vast majority of infectious disease outbreaks are only partially observed, with the proportion of missing cases being typically unknown as well (O’Neill and Roberts, 1999; Jewell et al., 2009; Chis Ster et al., 2009). These missing intermediates in the transmission trees represent a challenge for the integration of epidemiological data, since by definition there is no epidemiological data available on unknown putative hosts. This difficulty was noted, for example, when a spatial-genetic framework assuming complete sampling (Morelli et al., 2012) was extended to handle incomplete sampling, with two extreme scenarios proposed as bounds on the probability of spatial dispersion for unknown cases (Mollentze et al., 2014).

A naive approach to incorporate epidemiological data into the transmission analysis of a partially sampled outbreak is to consider all possible combinations for the epidemiological data of the unsampled cases along with their associated probabilities. For each combination the epidemiological component of the likelihood can be calculated as previously described in the case of a fully sampled outbreak (Ypma et al., 2012; Morelli et al., 2012; Didelot et al., 2014; Hall et al., 2015). The unconditioned likelihood is then be obtained as the average of these conditioned likelihoods, weighted according to their probabilities, using the law of total probability. However, the number of combinations scales exponentially with the number of unsampled cases in the transmission tree, and is only be computationally feasible for very small outbreaks. Another approach is to rely on data augmentation techniques within a Markov Chain

Monte-Carlo (MCMC) framework, in order to treat the epidemiological data of unsampled cases as additional parameters (van Dyk and Meng, 2001; O’Neill, 2002). Again this may not scale well to larger outbreaks, especially since the number of unsampled cases is unknown, so that efficient reversible jump proposals are required to deal with the transdimensional parameter space (Green, 1995; Sisson, 2005). Instead, we present a computationally efficient approach to calculate the epidemiological component of the likelihood. We show that this computation can be used to incorporate epidemiological data into the transmission analysis, boosting the accuracy of the analysis and generating the type of who-acquires-infection-from-whom matrices that are the cornerstone of predictive modelling. We illustrate our method on simulated datasets, before considering real-world examples of tuberculosis and H7N7 outbreaks.

### NEW APPROACHES

We take as our starting point the TransPhylo methodology (Didelot et al., 2014), which represents the transmission tree by colouring the branches of an input dated phylogeny (Rieux and Balloux, 2016). The first version of TransPhylo considered only fully sampled outbreaks, so that it was possible to incorporate epidemiological data (Didelot et al., 2014). With the extension of TransPhylo to the more generally useful situation of a partially sampled outbreak, fvthis possibility to integrate epidemiological data was lost (Didelot et al., 2017, 2021). More recently, TransPhylo was further extended to allow some hosts to be sampled more than once and to remove the assumption of complete transmission bottleneck (Carson et al., 2024), and this is the version that we use as our starting point for the incorporation of epidemiological data.

We extend the TransPhylo framework to incorporate known discrete epidemiological data on the sampled hosts, or a subset of them. Note that we use the term ‘deme’ to represent data that could be any discrete property of the hosts, for example geographical location in different towns or hospital wards, age or gender categories, classification based on behavioural data, infectious status from other infectious diseases, etc. We let *S* denote the number of demes (number of discrete epidemiological states). The transmission model within TransPhylo is a continuous time branching process (Farrington et al., 2003), in which each infected host generates a number of offspring *k* from an offspring distribution function *α*(*k*), and their infection times *τ* relative to the infection time of the infector are sampled from a generation time distribution *γ*(*τ*). The mean of the offspring distribution is the basic reproduction number *R*. We extend this branching process so that a deme is sampled for each offspring conditional on the deme of the infector. Specifically, the probability that a newly infected host belongs to deme *j* given that their infector belongs to deme *i* is denoted *P*_*ij*_. This matrix may take any form, as long as each of the rows sums up to one, and may include some parameters that we wish to infer jointly with the transmission tree.

We consider two complementary cases. In the first case, hosts in every deme have the same offspring distribution function and probability of being sampled. In this case the likelihood can be decomposed as the product of the transmission tree and of the epidemiological data, with the latter being calculated efficiently using a dynamic programming algorithm similar to the Felsenstein pruning algorithm (Felsenstein, 1973, 1981). In the second case, the offspring distribution function and probability of being sampled depend on the deme. This dependency may once again involve some parameters that we wish to estimate, for example different values *R*_1_, …, *R*_*S*_ for the basic reproduction number within each of the demes. In this case the likelihood can no longer be decomposed as previously, but we show that it can still be calculated analytically using a more complicated dynamic programming algorithm.

## RESULTS

### Exemplary analysis of a simulated dataset where all demes have the same offspring distribution and sampling probabilities

We simulate an outbreak with 250 observed infected hosts across five demes, with each observed host being sampled once. The observation cut-off time *T* is determined by the simulation in order to return the correct number of observed infected hosts. The generation time and primary observation time are both Gamma-distributed with shape and scale parameters equal to 2 and 1, respectively. For the transmission model, the offspring distribution follows a Negative Binomial distribution with *r* = 2 and *p* = 0.5, so the basic reproduction number *R* = *r* = 2, and the sampling proportion is *π* = 0.8. The within-host pathogen population size is *κ* + *λτ* at time *τ* after infection, with *κ* = 0.1 and *λ* = 0.2. The probability of an offspring having the same deme as their infector is *ρ* = 0.8, otherwise one of the other four demes is sampled uniformly. The resulting simulation contains 302 infected hosts (of which 250 are sampled). The transmission and phylogenetic trees are shown in Figure 1, which is coloured according to the demes of the hosts. Note that only the deme data for the observed hosts is used in the analysis.

**Figure 1:**
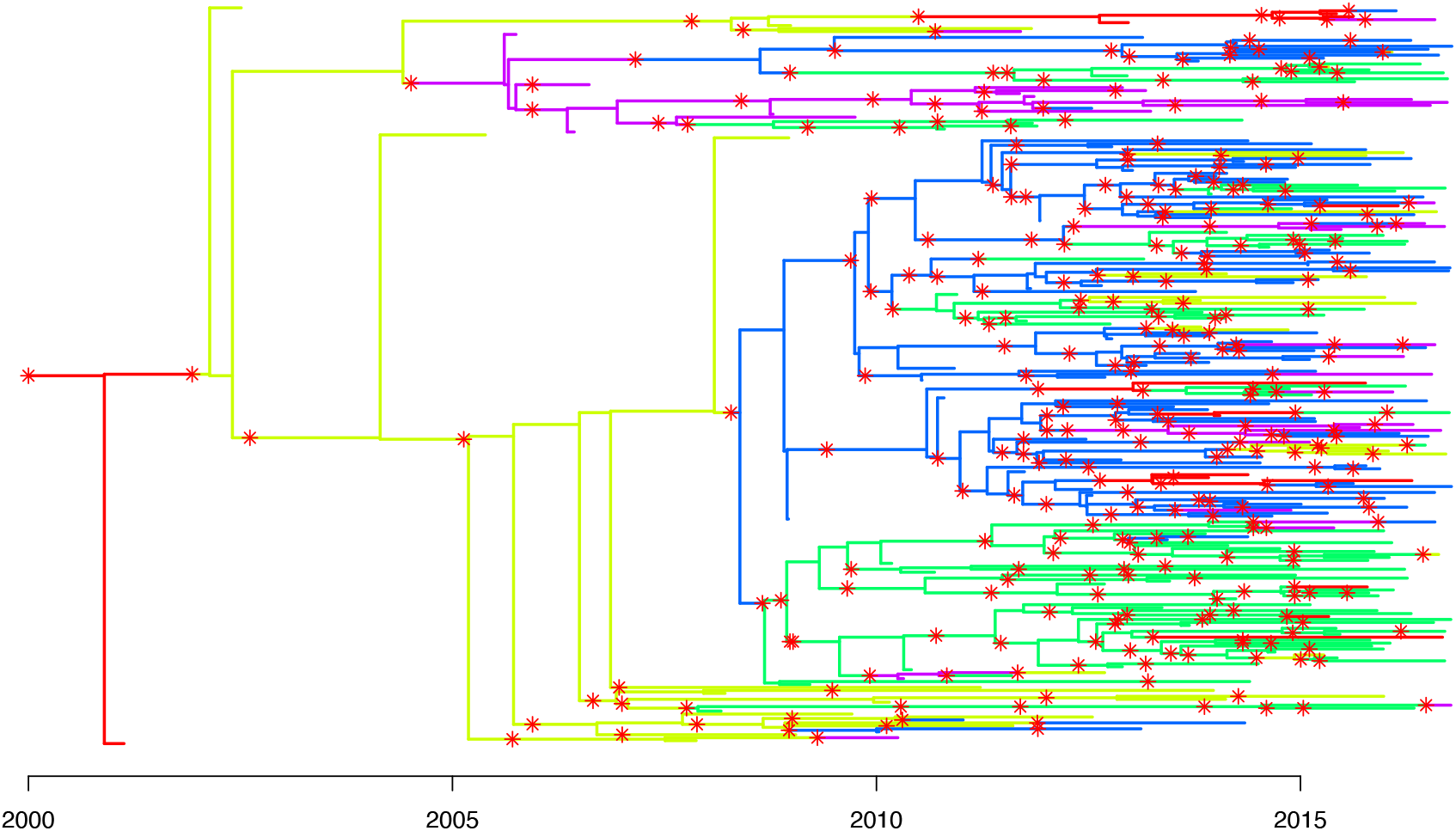
Combined transmission and phylogenetic tree coloured by host deme used in the first simulation study. The tree contains 302 infected hosts, of which 250 are sampled. The red stars correspond to transmission events.

Our goals are to estimate the five parameters *R, π, ρ, κ* and *λ*, and to correctly identify transmission links between sampled hosts. We performed four separate MCMC runs of 100,000 iterations, which each took approximately 36 hours on a 3 GHz processor core. Mixing was relatively slow for the coalescent parameters *κ* and *λ*, with effective sample sizes of 300-600 in each run. This is in part due to these parameters being highly correlated to the transmission tree, which is updated separately within the MCMC algorithm, and in part due to having only one sample per host, leading to a wide posterior to explore for these parameters (Carson et al., 2024). The effective sample size was between 1300-1900 in each chain for *π*, 6000-7000 for *R*, and 6000-10000 for *ρ*. The multivariate Gelman-Rubin statistic comparing runs was 1.01 (Brooks and Gelman, 1998). The inferred means (95% credible intervals) for each parameter are *R* : 1.94 (1.68, 2.22), *π* : 0.78 (0.63, 0.93), *ρ* : 0.78 (0.73, 0.83), *κ* : 0.09 (0.00, 0.21), *λ* : 0.20 (0.02, 0.41). This shows that we are able to recover the simulated parameter values effectively, since the posterior means are close to the correct values and the credible intervals cover the correct values.

In order to evaluate our ability to reconstruct transmission links, we focus on transmissions between observed hosts. Out of the 250 observed hosts, 184 are infected by another observed host. The posterior probability estimates for the transmission links are summarised in Figure 2. If we define 0.5 as the posterior probability threshold for a transmission event being identified, we correctly identify 71 transmission links including the direction of transmission, giving a directional sensitivity of 39%. With only one observation per host it is common to identify a transmission link between two hosts, but be unsure of the direction of transmission (Didelot et al., 2014, 2017; Carson et al., 2024). If we ignore the direction of transmission we identify 104 transmission links, giving a bidirectional sensitivity of 57%. We incorrectly establish 30 directional transmission links, and 37 bidirectional transmission links. However, as there are 62,250 possible host combinations, specificity is high (*>* 99.9%) in both cases. The resulting precision is 70% when including the direction of transmission, and 74% when ignoring the direction of transmission.

**Figure 2:**
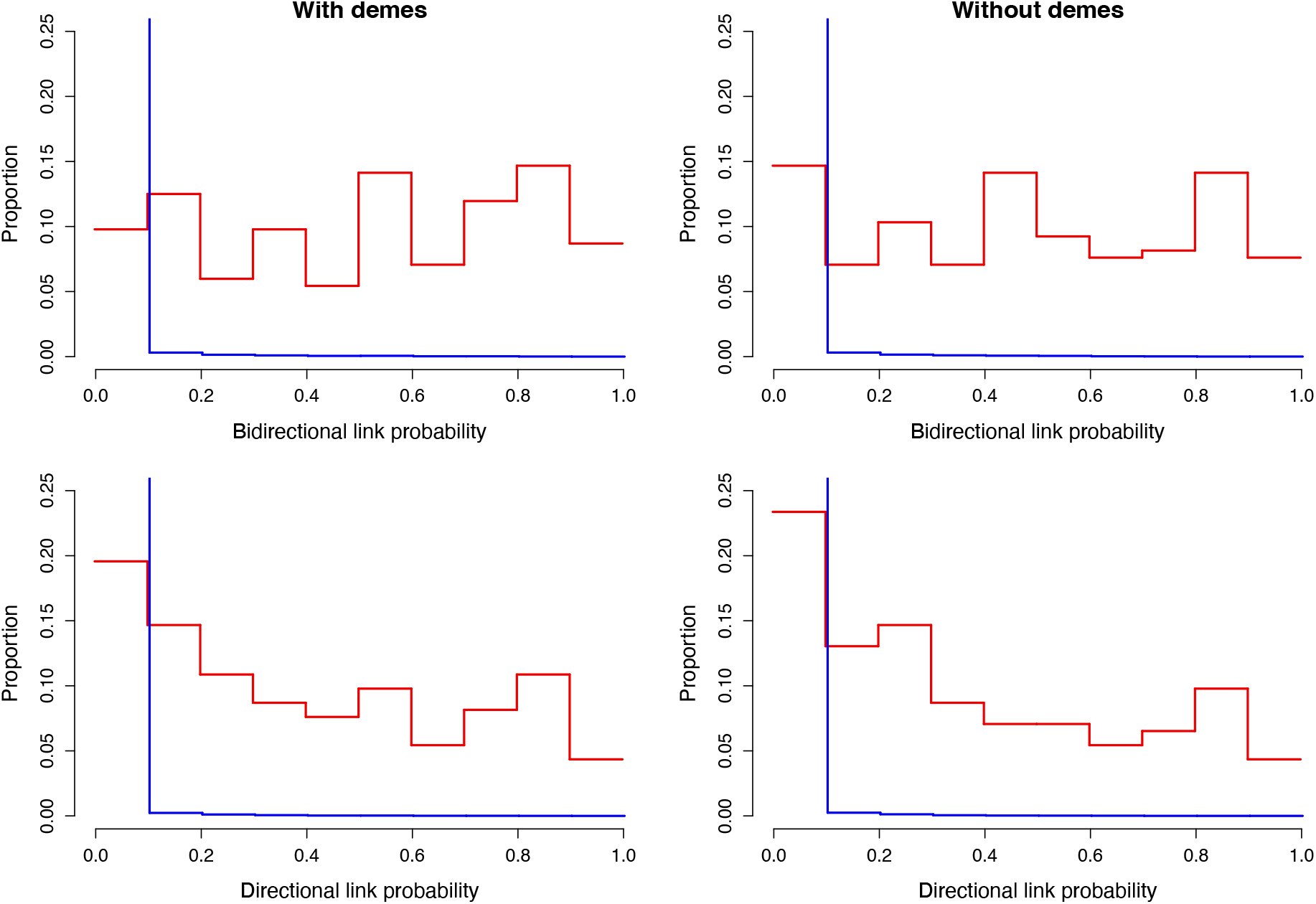
Posterior probability estimates for the transmission links in the first simulation study. The red lines correspond to correct transmission links, and the blue lines correspond to incorrect transmission links. In each case the posterior probability estimates are binned in 0.1 width intervals, and the vertical height indicates the proportion of transmission links contained within each bin. The left plots show estimates with deme information, and the right plots show estimates without deme information. The top plots show bidirectional transmission link estimates, and the bottom plots show directional transmission link estimates.

If inference is undertaken without the deme data we obtain similar parameter estimates for *R, π, κ*, and *λ*, but no longer obtain an estimate for *ρ* as we assume that all hosts belong to a single deme. We correctly identify fewer transmission links, with 61 correct links being established if we consider direction (directional sensitivity of 33%) and 86 if we do not (bidirectional sensitivity of 47%). However, we also obtain a slightly smaller number of false positives: 25 in the directional case and 32 in the bidirectional case. The resulting precision changes very little, with a precision of 71% when including the direction of transmission, and 73% otherwise.

### Exemplary analysis of a simulated dataset where demes have different offspring distributions and sampling probabilities

We simulate a second outbreak with 250 observed hosts. The hosts belong to two demes, with each deme having its own *R, π*, and *ρ* parameters. Specifically we set *R*_1_ = 1.2, *R*_2_ = 2.2, *π*_1_ = 0.4, *π*_2_ = 0.9, *ρ*_1_ = 0.9, *ρ*_2_ = 0.7, whilst maintaining *κ* = 0.1 and *λ* = 0.2. Consequently, deme 1 has a low transmission rate and is poorly surveyed, whilst deme 2 has a high transmission rate and is well surveyed. Offspring are more likely than not to be in the same deme as the infecting host, but transmissions from deme 2 to deme 1 are more likely than transmissions from deme 1 to deme 2. The resulting simulation contains 325 hosts, and the transmission and phylogenetic trees are shown in Figure 3, which is coloured according to the demes.

**Figure 3:**
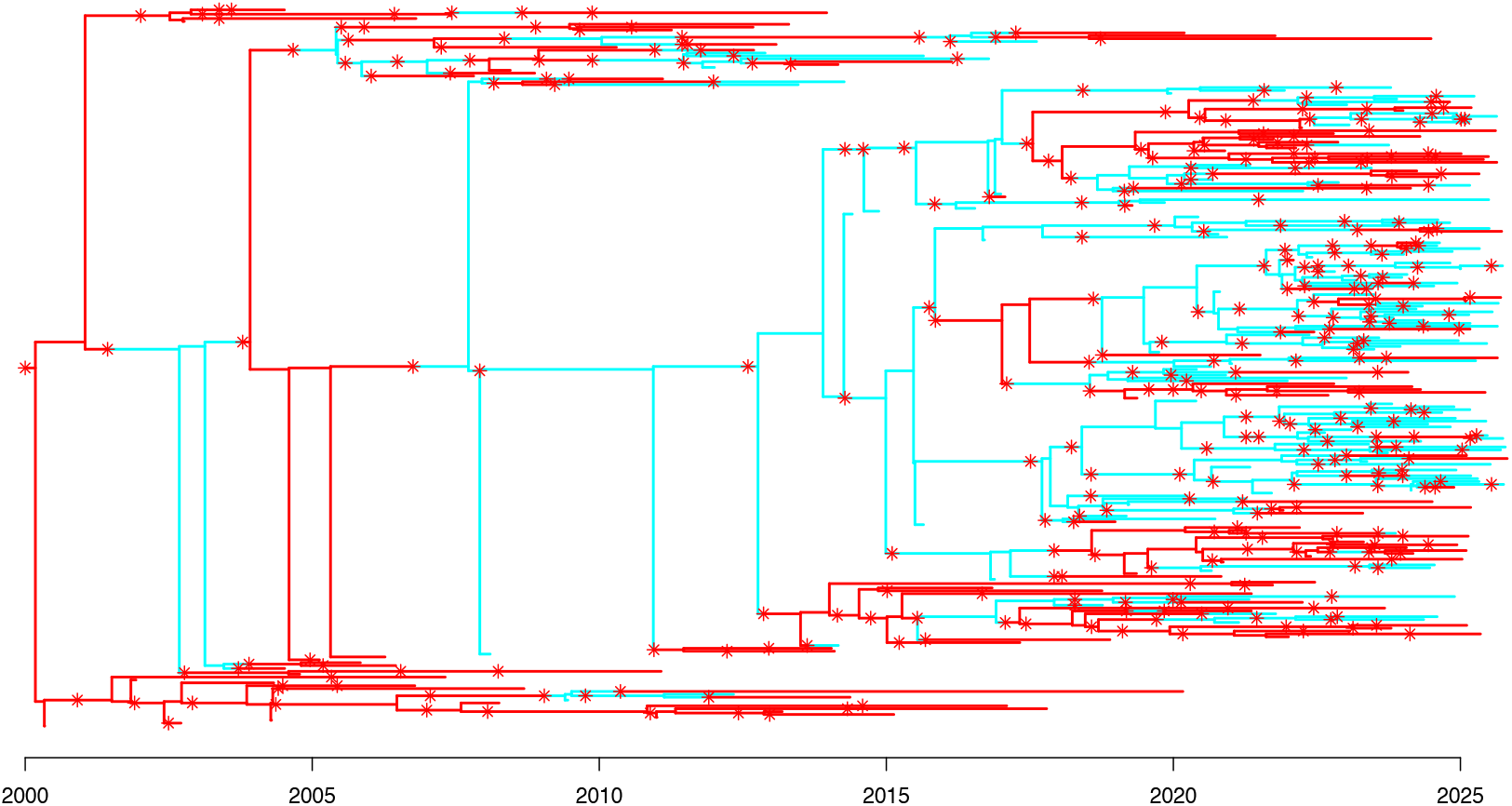
Combined transmission and phylogenetic tree coloured by host deme used in the second simulation study. The tree contains 325 infected hosts, of which 250 are sampled. The red stars correspond to transmission events.

We performed four separate MCMC runs of 100,000 iterations, which took approximately 46 hours on a 3 GHz processor core. As with the previous simulation study, mixing was slowest for *κ* and *λ*, with effective sample sizes between 300-1100 for *κ* and 300-400 for *λ* in each chain. For the remaining parameters the effective sample sizes were 4000-5500 for *R*_1_, 500-4200 for *R*_2_, 2000-2300 for *π*_1_, 1700-2300 for *π*_2_, 900-3700 for *ρ*_1_, and 1500-3300 for *ρ*_2_. The multivariate Gelman-Rubin statistic comparing runs was 1.01 (Brooks and Gelman, 1998). The inferred means (and 95% credible intervals) for each parameter are *R*_1_ : 1.15 (0.92, 1.39), *R*_2_ : 2.21 (1.78, 2.68), *π*_1_ : 0.50 (0.35, 0.68), *π*_2_ : 0.88 (0.68, 0.99), *ρ*_1_ : 0.93 (0.87, 0.96), *ρ*_2_ : 0.72 (0.62, 0.81), *κ* : 0.10 (0.00, 0.28), *λ* : 0.23 (0.03, 0.48). Once again this shows that we are able to recover the simulated parameter values effectively since the inferred values are close to the correct values used in the simulation. Furthermore, since the credible intervals for *R*_1_ and *R*_2_, and for *π*_1_ and *π*_2_ do not overlap, we can deduce that we have correctly inferred that the reproduction number and sampling probability are both higher in the second deme than in the first deme.

Looking once more at inferred transmission links, out of the 250 observed hosts 155 are infected by another observed host. The full transmission link probabilities are shown in Figure 4. Using the same posterior probability threshold as the first simulation study we correctly identify 65 transmission links including the direction of transmission, giving a directional sensitivity of 42%. If we ignore the direction of transmission then we identify 89 transmission links, giving a bidirectional sensitivity of 57%. We incorrectly establish 39 directional transmission links, and 50 bidirectional transmission links. These values indicate a precision of 63% when including the direction of transmission, and 64% otherwise.

**Figure 4:**
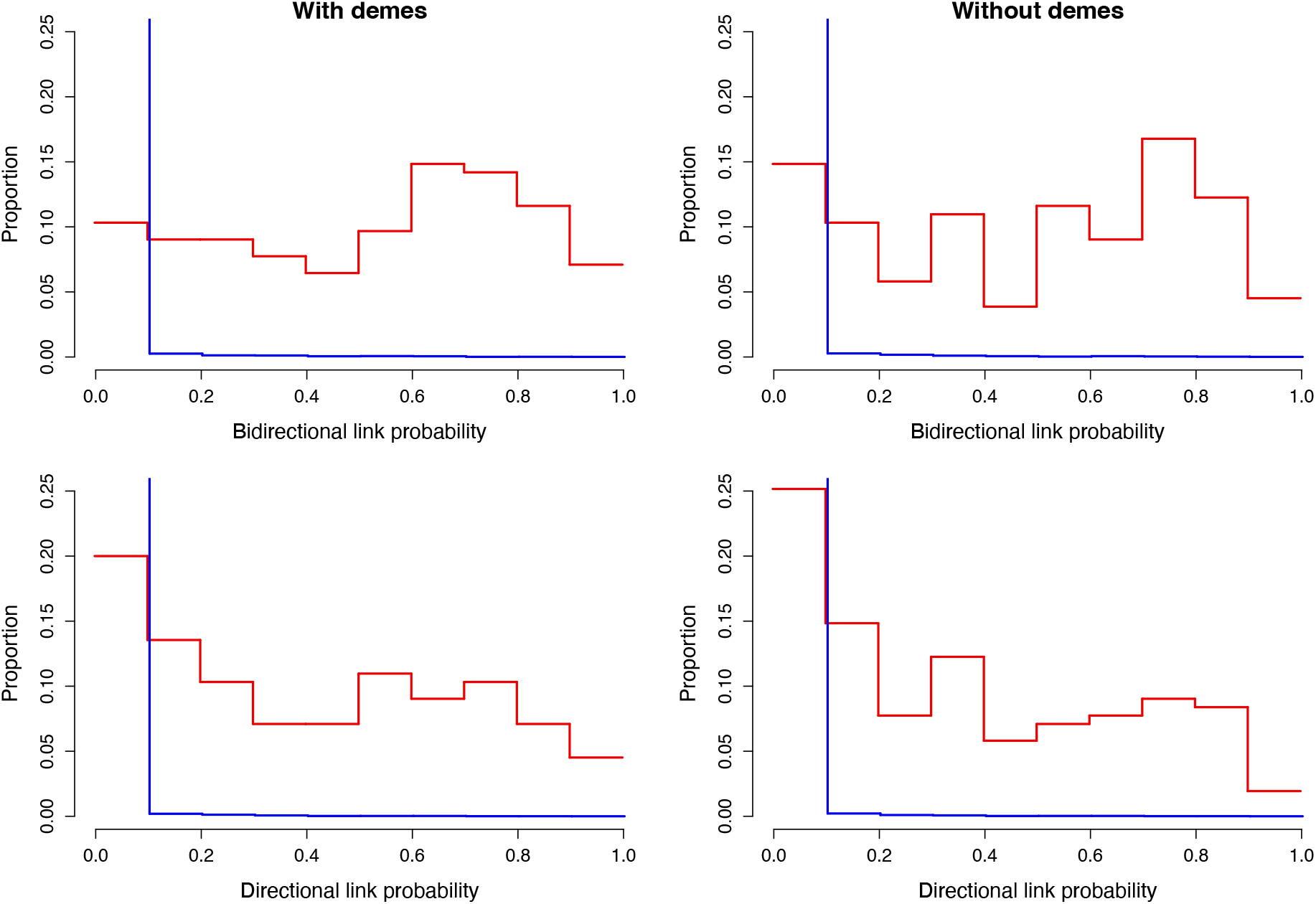
Posterior probability estimates for the transmission links in the second simulation study. The red lines correspond to correct transmission links, and the blue lines correspond to incorrect transmission links. In each case the posterior probability estimates are binned in 0.1 width intervals, and the vertical height indicates the proportion of transmission links contained within each bin. The left plots show estimates with deme information, and the right plots show estimates without deme information. The top plots show bidirectional transmission link estimates, and the bottom plots show directional transmission link estimates..

When undertaking inference without deme data we assume that all hosts belong to the same deme, giving single *R* and *π* parameters, and removing all *ρ* parameters. For the posterior means and credible intervals we find *R* : 1.56 (1.35, 1.79), *π* : 0.73 (0.58, 0.9), *κ* : 0.9 (0.00, 0.25), and *λ* : 0.28 (0.04, 0.58). That is, we estimate *R* and *π* as somewhere between the pairs of values used in the simulation. The true values of *κ* and *λ* remain in the 95% credible intervals, but the estimate of *λ* has slightly increased. In this instance we identify fewer correct transmission links, and more incorrect transmission links. Specifically, 53 links are established if we consider direction (directional sensitivity of 34%) and 84 if we do not (bidirectional sensitivity of 54%). We find 47 false positives in the directional case and 50 in the bidirectional case. This means that the precision is significantly smaller when including the direction of transmission (53%), but similar when ignoring the direction of transmission (63%).

### Benchmarking using multiple simulations

We undertake 50 simulation studies across a range of parameter values, similar in design to the second simulation study. The parameter sets are sampled from an orthogonal array Latin hypercube using the lhs R package. In each case we simulate an outbreak with 250 observed hosts across two demes, with each observed host being sampled once. Each deme has separate *R* values sampled between 1 and 6, separate *π* values sampled between 0.1 and 1, and separate *ρ* values sampled between 0.5 and 0.9. Smaller values of *ρ* are not considered, as if *ρ* is high in one deme and low in the other this will lead to samples being dominated by one deme. In such cases we would expect to obtain poor parameter estimates for the less sampled deme. The remaining parameters are the same for both demes, namely both *κ* and *λ* are sampled between 0 and 1. For each simulated dataset we estimate the eight parameters used in the simulation.

The marginal posterior results credible intervals are shown in Figure 5 and compared to the correct values of the parameters used in the simulations. In general we are able to recover the parameter values used in each simulation, but there is considerable uncertainty on some of the parameters, as can be seen by the wide credible intervals in Figure 5. The uncertainty is particular high for parameters of a deme with a small number of representative samples, as shown by the more lightly shaded bars being longer than the more darkly shaded bars in Figures 5A-F. The parameter *ρ* tends to be inferred more precisely when its correct value is high (Figure 5E-F. The parameters *λ* and *κ* of the within-host population size are only weakly informed (Figures 5G-H) as previously noted in the two exemplary analyses.

**Figure 5:**
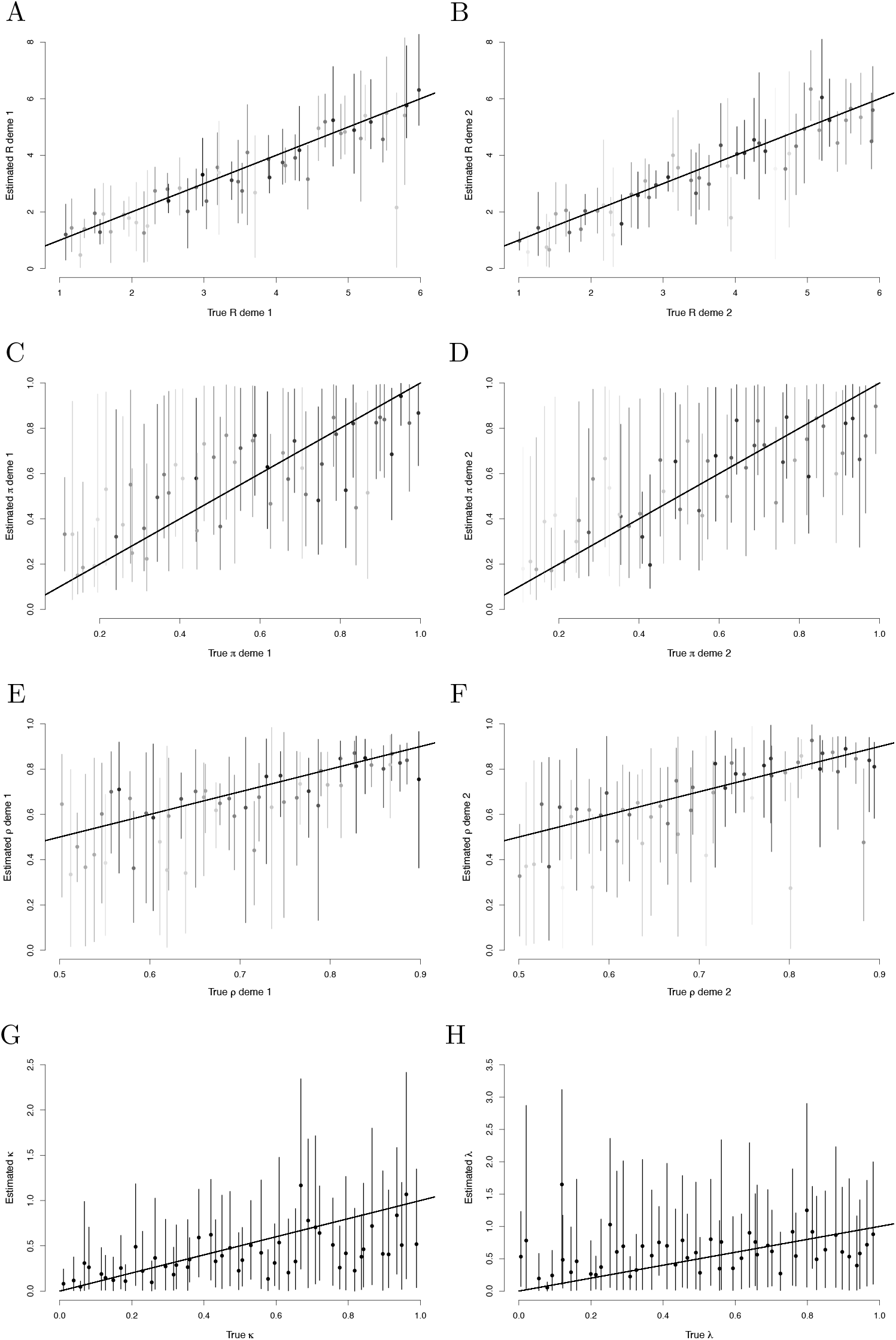
Benchmarking results for the parameters *R* of deme 1 (A), *R* of deme 2 (B), *π* of deme 1 (C), *π* of deme 2 (D), *ρ* of deme 1 (E), *ρ* of deme 2 (F), *κ* (G) and *λ* (H). Mean values are shown by dots, 95% credible intervals are shown by vertical lines. In A-F the shade indicates the proportion of sampled hosts in the associated deme (darker implying a greater proportion in the deme).

Averaging across the 50 simulated data sets, 106.8 out 250 observed hosts are infected by another sampled host. Using a posterior probability threshold of 0.5, on average we correctly identify 15.2 transmission links including the direction of transmission, and 26.6 transmission links ignoring the direction of transmission. These correspond to an average directional sensitivity of 14%, and an average bidirectional sensitivity of 24%. These relatively low sensitivities are typical of data sets with one sample per observed host, and data sets with a relaxed bottleneck (Carson et al., 2024). On average we also find 9.3 false positives including the direction of transmission, and 14.1 false positives when ignoring the direction of transmission. The corresponding directional and bidirectional specificities are *>* 99.9%. The average directional precision is 61%, and the average bidirectional precision is 64%.

We obtain the sensitivity (recall), specificity, and precision for each simulation under a series of different posterior probability thresholds between 0 and 1. Averaging over the 50 simulated datasets, we present the resulting Receiver Operating Characteristic (ROC) and Precision-Recall (PR) curves in Figure S1 for both directional and bidirectional transmission links. The Area Under the Curve (AUC) is 0.99 for both ROC curves, 0.33 for the PR curve of directional transmission links, and 0.44 for the PR curve of bidirectional transmission links.

### Application to an outbreak of tuberculosis in Argentina

A multidrug resistant *Mycobacterium tuberculosis* outbreak in Argentina has been described and studied in detail using genomic epidemiology (Eldholm et al., 2015). A total of 252 genomes were sequenced, with collection dates ranging between 1996 and 2010. 153 of the genomes originated from HIV positive individuals, whereas the remaining 99 genomes where sampled from HIV negative individuals. A dated phylogeny was previously reconstructed using BEAST (Drummond et al., 2012), with the root of this tree being estimated to have existed around 1970 (Eldholm et al., 2015). This dated phylogeny is the starting point of our analysis and reproduced in Figure S2, with leaves colored according to the HIV status of the hosts. The role of HIV co-infection in the transmission of this tuberculosis outbreak has been previously investigated and found to be not statistically significant (Eldholm et al., 2016). However, this analysis was based on a rough reconstruction of transmission events, with a-posteriori testing of the effect of HIV status, limiting its statistical power (Eldholm et al., 2016). It is therefore interesting to reanalyse this dataset with the new methodology presented here, considering two demes for the HIV positive and negative individuals. The sampling window was set from 1st October 1996 to 1st December 2009 to include all samples and reflect the original sampling collection methodology (Eldholm et al., 2015). We used the same generation time and sampling time distributions as was used in previous analyses of tuberculosis outbreaks (Didelot et al., 2017; Séraphin et al., 2018; Sobkowiak et al., 2023; Chitwood et al., 2024).

The means (95% credible intervals) of the parameters of the within-host population size function are *κ* : 5.38 (2.82, 8.79) and *λ* : 0.63 (0.04, 1.69). The initial pathogen population size *κ* is large compared to the per-year linear growth rate *λ*, suggesting a relaxed transmission bottleneck (Carson et al., 2024). Figure 6 shows the posterior distribution for the parameters specific to both demes. The reproduction number *R* for the HIV negative and HIV positive demes are 1.10 (0.65, 1.52) and 1.63 (0.86, 2.32), respectively (Figure 6A). The probability that the HIV positive deme has a greater reproduction number than the HIV negative deme is 0.86. The sampling probability *π* for the HIV negative and HIV positive demes are 0.32 (0.13, 0.70) and 0.79 (0.46, 0.99), respectively (Figure 6B). The probability that the HIV positive deme has a greater sampling probability than the HIV negative deme is 0.98. These results suggest that HIV positive individuals have a greater reproduction number and are more likely to be observed, as would be expected from the fact that HIV co-infection accelerates the transition from latent to active tuberculosis (Bruchfeld et al., 2015).

**Figure 6:**
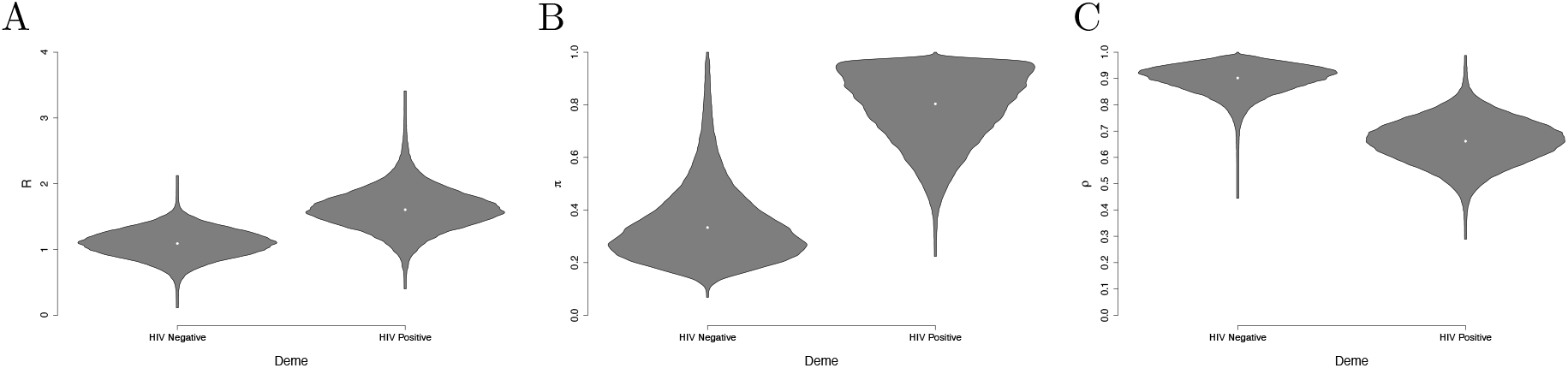
Parameter estimates in the tuberculosis analysis: the reproduction number *R* (A), sampling fraction *π* (B) and probability to remain in a deme *ρ* (C) are shown for both the HIV negative (left) and HIV positive (right) demes.

Figure 7 shows the posterior probabilities of direct transmission from any individual to any other. It is visually clear that infector/infectee pairs tend to have the same HIV status. This is confirmed by the estimate of the probabilities that the pathogen remains in the same deme at transmission which are 0.90 (0.77, 0.98) and 0.66 (0.48, 0.82), respectively for the HIV negative and HIV positive demes (Figure 6C). Therefore HIV negative hosts are highly likely to transmit to other HIV negative hosts. HIV positive hosts are also more likely to transmit to other HIV positive hosts, but to a lesser extent. This may be in part due to the HIV negative population being larger than the HIV positive population.

**Figure 7:**
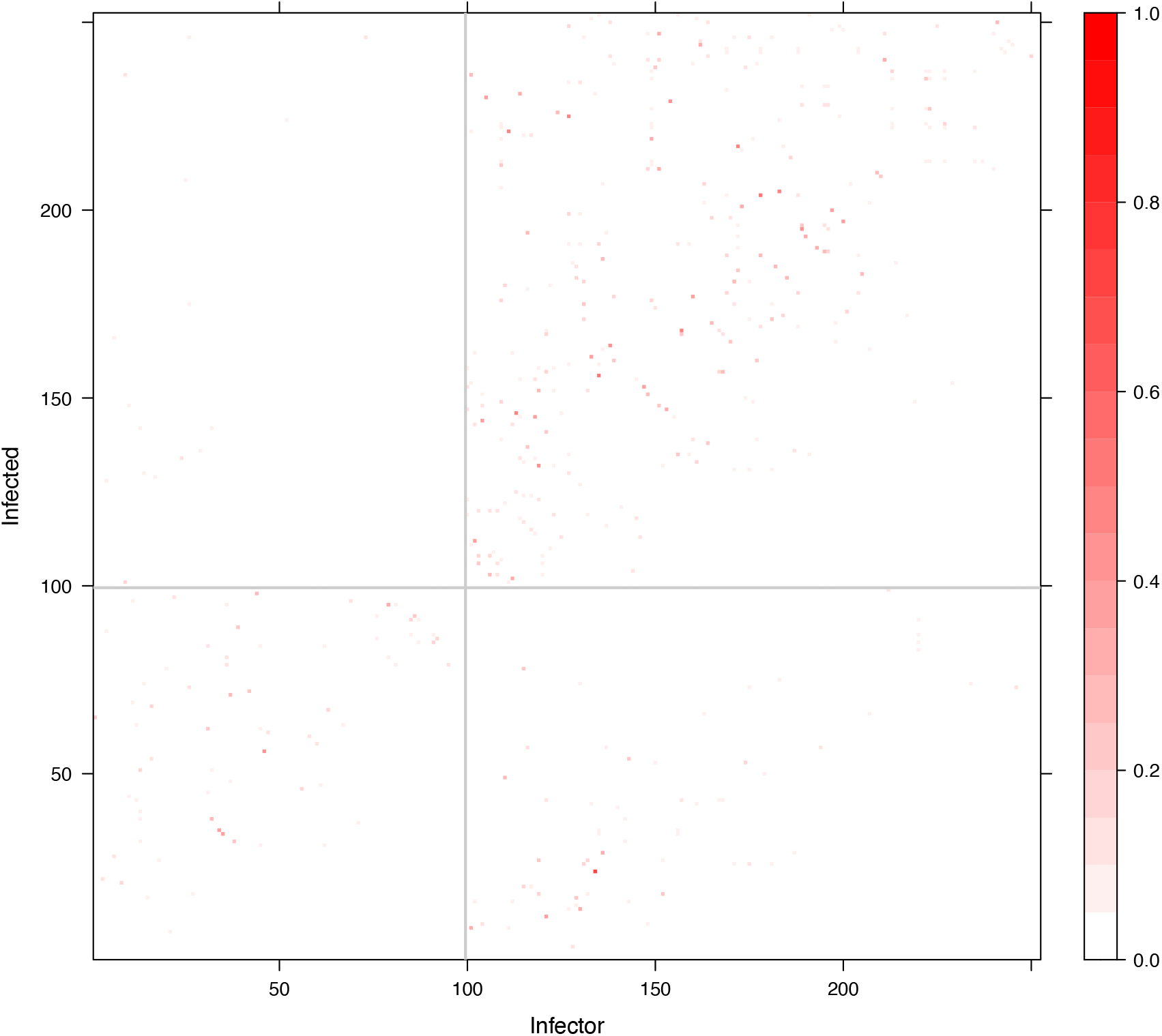
Posterior direct transmission probabilities in the tuberculosis analysis. The grey lines separate the two demes, with HIV negative being left/bottom and HIV positive being right/top.

### Application to an outbreak of avian flu H7N7 outbreak in the Netherlands

An outbreak of avian influenza H7N7 occurred in the Netherlands during 2003, infecting 255 Dutch farms in less than 3 months, and leading to drastic control measures including the culling of 30 million birds (Stegeman et al., 2004). Genetic data is available from GISAID (Shu and McCauley, 2017) from 227 farms, for genes HA, NA and PB2 which were concatenated. Most sequences are from the Gelderland (G) area (*n* = 186) with smaller numbers from the Limburg (L) area (*n* = 33), Central (C) area (*n* = 7) and Southwest (S) area (*n* = 1). The phylogeography of this outbreak has been described before in a number of studies (Bataille et al., 2011; Ypma et al., 2012, 2013; Hall et al., 2015; Klinkenberg et al., 2017). We built a dated tree using BEAST2 (Bouckaert et al., 2019) which is shown in Figure S3 with leaves colored by location. We used the same generation time and sampling time distributions as in a recent study of this outbreak (Klinkenberg et al., 2017). The sampling window was set from the 50th to the 125th day from the root of the dated tree, which included all samples (Figure S3).

The means (95% credible intervals) of the parameters of the within-host population size function are *κ* : 5.38 (2.23, 9.60) and *λ* : 3.55 (1.24, 6.28). Figure 8 shows the posterior distribution for the parameters specific to the four locations. The per-location reproduction numbers are 1.00 (0.86, 1.16), 0.98 (0.71, 1.25), 1.13 (0.54, 1.93) and 0.74 (0.02, 2.37) for regions G, L, C and S, respectively. These reproduction numbers are close to 1, with uncertainty increasing as the sample numbers decrease (Figure 8A). In location S we approximately recover the prior exponential with mean 1, as would be expected given that there was only a single representative of this location. The sampling probabilities *π* are 0.71 (0.36, 0.97), 0.28 (0.10, 0.55), 0.33 (0.07, 0.82), 0.27 (0.02, 0.83) for regions G, L, C and S, respectively. Location G is best sampled, which makes sense given that it has the largest number of sampled cases (Figure 8B). The remaining demes are likely less well sampled, although there was high uncertainty due to the small sample numbers. Our analysis was conducted on 227 farms for which genetic data was available, whereas during the outbreak 255 farms were confirmed to be infected Stegeman et al. (2004); Bataille et al. (2011), suggesting an upper bound of the sampling fraction of 0.89. Our estimates are compatible with this, and further suggest that some infection went undetected as would be expected from the large scale culling that took place at farms even in absence of detection Stegeman et al. (2004).

**Figure 8:**
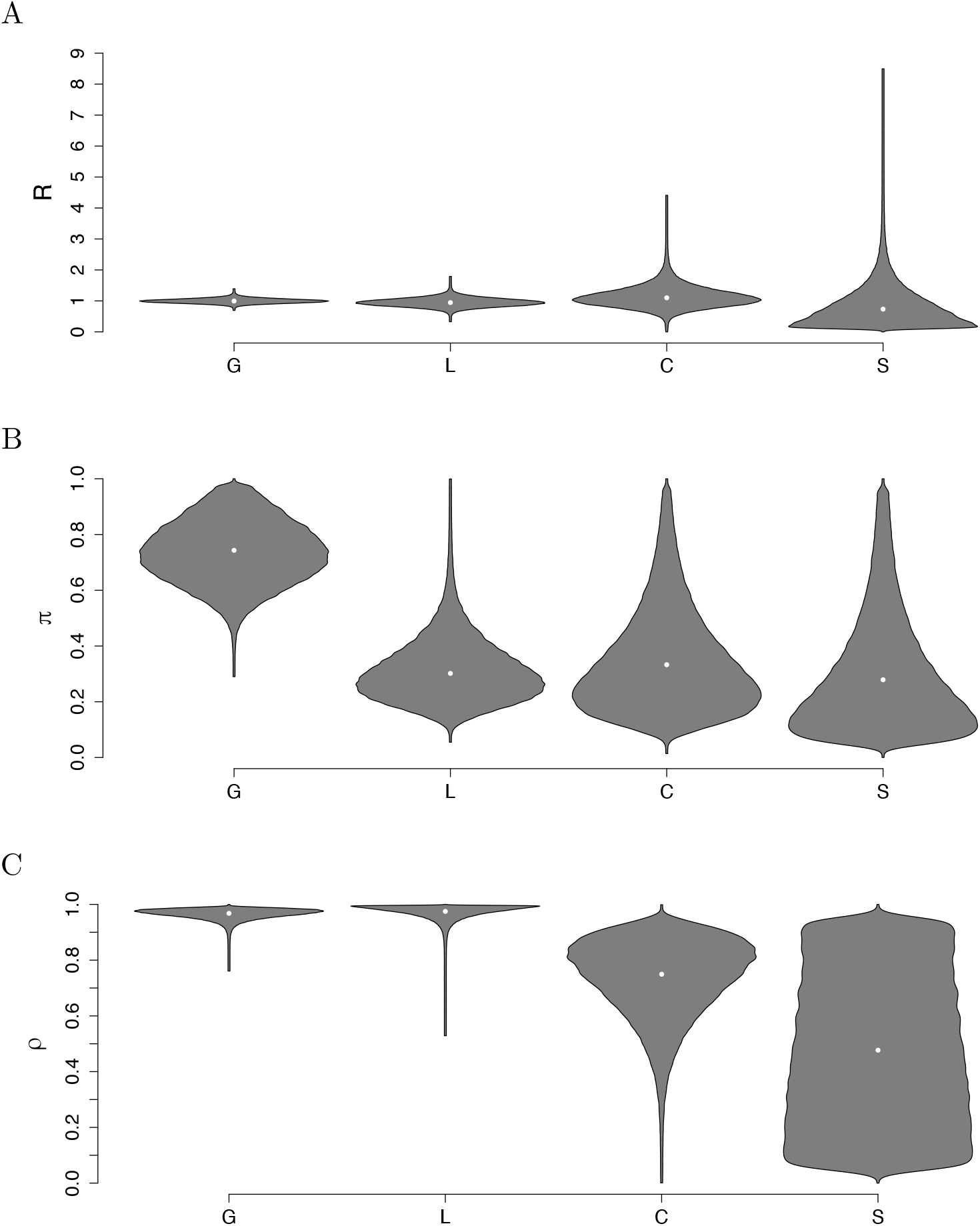
Parameter estimates in the H7N7 analysis: the reproduction number *R* (A), sampling proportion *π* (B) and probability to remain in a deme *ρ* (C) are shown for each of the four locations.

Figure 9 shows the posterior probabilities of direct transmission from any farm to any other, with a clear tendency for farms to infect other farms from the same region. This is confirmed by the parameters *ρ* representing the probability that the pathogen infects in the same location which are estimated to be 0.97 (0.92, 0.99), 0.98 (0.92, 1.00), 0.75 (0.4, 0.95) and 0.48 (0.02, 0.97) for regions G, L, C and S, respectively (Figure 8C). Note that we assume that transmissions to different locations are evenly distributed across the other locations. Farms in locations G and L almost always transmit to offspring in the same deme. The probability that a farm in location C infects another farms in location C also seems high, but is more uncertain due to the small number of samples. For location S represented by only a single farm we approximately recover the prior uniform between 0 and 1.

**Figure 9:**
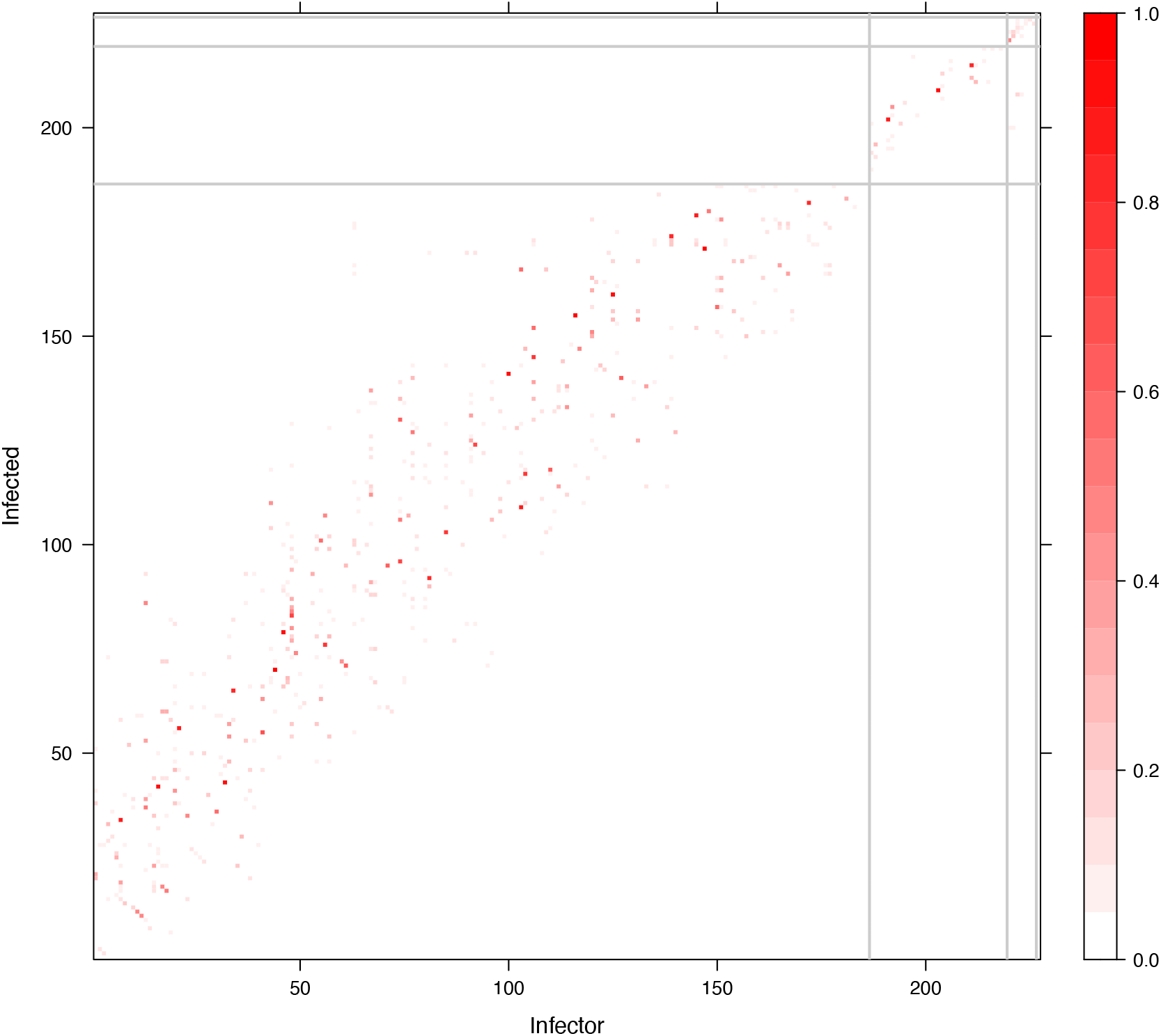
Posterior direct transmission probabilities in the H7N7 analysis. The grey lines separate the four demes.

## DISCUSSION

Combining genomic and epidemiological data to reconstruct the transmission events within an infectious disease epidemic is an idea that was formulated over a decade ago, when the first methods to use genomic data for outbreak reconstruction were proposed (Ypma et al., 2012; Jombart et al., 2014; Didelot et al., 2014; Hall et al., 2015). It is however difficult to use epidemiological data when considering partially sampled or ongoing outbreaks (Mollentze et al., 2014; Didelot et al., 2017), since unsampled cases do not have any associated epidemiological data. A naive approach to this issue quickly becomes intractable as larger numbers of unsampled cases need to be considered. Instead we presented a new dynamic programming algorithm that can efficiently resolve this problem. We implemented this new methodology by extending the TransPhylo framework which can be applied to partially sampled outbreaks (Didelot et al., 2017, 2021), even when multiple genomes per host are provided or when the transmission bottleneck is not complete (Carson et al., 2024).

We used simulations to show that the combined approach has improved statistical power to infer the correct transmission events compared to the previous approach based on genomic data only. We also showed that the parameters governing the epidemiological data can be inferred with accuracy that increases with the amount of data available for analysis. We applied our new algorithm to two widely different real datasets to showcase the range of scenarios in which it can be useful. First we analysed data from a tuberculosis outbreak in Argentina (Eldholm et al., 2015) to investigate the role of HIV co-infection on the spread of the bacterial causative agent *M. tuberculosis*. Second we analysed data from the avian influenza H7N7 epidemic that hit the Netherlands in 2003 (Stegeman et al., 2004), to infer the parameters involved in the spatial spread of this virus from farm to farm.

The methodological framework we developed makes few assumptions, and we therefore envisage that it can be useful in a wide range of situations. The epidemiological model at the heart of TransPhylo is a flexible branching process (Didelot et al., 2017) based on offspring distribution and generation time distribution whose parameters can be set to appropriately model many infectious diseases transmitted directly from host to host (Wallinga and Teunis, 2004; Grassly and Fraser, 2008; Cori et al., 2013). An example of application concerns the inference of the different reproduction numbers for different components of the population. This can help determine their relative contribution to the overall disease burden, and therefore inform how to target public heath policies for maximum effect (Fraser et al., 2004; Grassly and Fraser, 2008; Hollingsworth, 2009). Another application likely to be useful is to estimate the sampling proportions for different components of the population, which can reveal if sampling is currently biased and how it could be improved (Magnani et al., 2005; Brooks-Pollock et al., 2021; Layan et al., 2023).

There are however some limitations to the methodology we presented. First, the analysis is based on a dated phylogeny that needs to be correctly precomputed. This requires consideration of how such a tree is computed, under which prior model if a Bayesian method is used, and to what extent a single point estimate can be used without quantification of uncertainty. These questions arise for all of the many recently developed phylodynamic methods that take a dated tree as input (Didelot and Parkhill, 2022). However, this step-by-step approach is necessary to be able to analyse state-of-the-art large genomic datasets. Second, we only considered discrete epidemiological data. Continuous variables can always be discretised to circumvent this limitation, but doing so may lose some information and requires to define potentially arbitrary discrete classes. This situation is analogous to the almost ubiquitous use of discrete locations in phylogeography (Lemey et al., 2009; De Maio et al., 2015; Baele et al., 2017). In our applications, we also assume that transmission outside of a deme is evenly distributed across the other demes. We make this choice in order to restrict the number of parameters to linear in the number of demes instead of quadratic, but recognise that it will not be appropriate for every situation. The dynamic programming algorithm allows for this assumption to be relaxed, but a more complex MCMC algorithm would be required to efficiently estimate the additional model parameters, and these estimates would likely exhibit greater uncertainty. Finally it should be noted that our methodology has a non-negligible computational cost. For example, the largest analysis we performed, on the H7N7 dataset, took several days to achieve acceptable MCMC convergence and mixing properties. However, our algorithm currently runs only on a single CPU core, whereas most standard desktop and laptop computers have 8 to 16 cores, with many more cores available on servers dedicated to computer-intensive tasks Future work should therefore seek to exploit multiple cores to reduce the overall runtime, for example by following recent progress in parallel MCMC algorithms (Schwedes and Calderhead, 2021; Syed et al., 2022; Glatt-Holtz et al., 2024).

## METHODS

### Case where all demes have the same offspring distribution and sampling probabilities

Let us start with the simpler case where all demes are assumed to have the same offspring distribution and sampling probabilities. In this case most of the calculations in TransPhylo (Didelot et al., 2017; Carson et al., 2024) remain unchanged, we simply include an additional likelihood term obtained by using an efficient dynamic programming algorithm similar to Felsenstein’s tree-pruning algorithm (Felsenstein, 1973, 1981). This is necessary to integrate over demes for unsampled individuals that form part of the transmission tree and for sampled individuals with missing deme data.

Let hosts be labelled 1, …, *N*, and define the deme of host *n* by *s*^*n*^ ∈ *{*1, …, *S}*. Let *P*_*ij*_ be the probability that an offspring of a host in deme *i* is in deme *j*. Finally, let 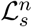 be the likelihood from the deme data of host *n* and their descendants, conditional on host *n* being in deme *s*.

The algorithm is initialised at the leaf nodes, which in this case are hosts with no offspring in the transmission tree, noting that all such hosts must have been sampled in TransPhylo. If the deme of such a host is known, then 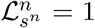 at the hosts deme *s*^*n*^, and 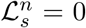 for *s /*= *s*^*n*^ (all other demes). If the deme of the host is unknown, then 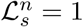 for all possible demes *s*.

The algorithm then proceeds backwards in time to evaluate the conditional likelihoods of the internal nodes (hosts with offspring). Let ℋ^*n*^ denote the set of offspring of Host *n*. If Host *n* has a known deme *s*^*n*^ then

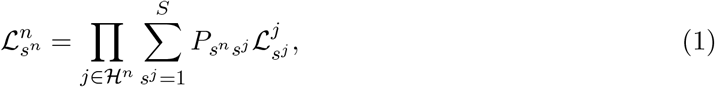

and 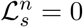 for *s /*= *s*^*n*^. If on the other hand Host *n* has no known deme then

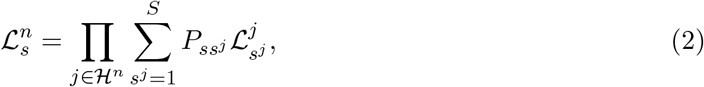

for all possible values of *s*.

The algorithm terminates at the root host, assumed here to be *n* = 1. The overall likelihood of the demes on the transmission tree is given by

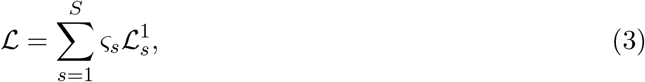

where *?*_*s*_ is the prior probability of the root host being in deme *s*.

### Illustrative example

An example of the dynamic programming algorithm is shown in Figure S4A. The target transmission tree contains 10 hosts and we assume that there are three possible demes. The transition probability between the three demes is given by

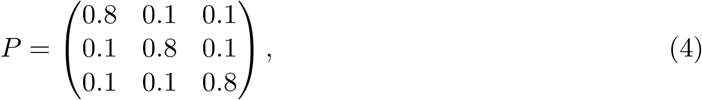

meaning that a host has probability 0.8 of an offspring having the same deme as its infector, and a probability 0.1 of an offspring being in either of the other two demes. We know that Host 5 is in deme 1, Hosts 3 and 7 are in deme 2, and Host 10 is in deme 3. Hosts 2, 4, and 9 are sampled hosts, but their deme is missing. Hosts 1, 6 and 8 are unsampled hosts.

Each host has an associated vector for the conditional likelihood at the three demes. Working backwards in time, Host 10 is known to be in deme 3 and is a leaf, and so:

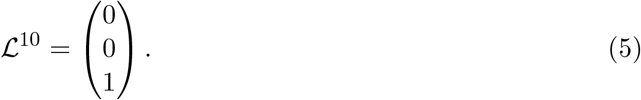

Host 9 is a leaf, but does not have a known deme, so that:

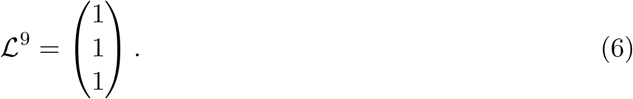

Host 8 is an unsampled individual, whose only offspring is Host 10:

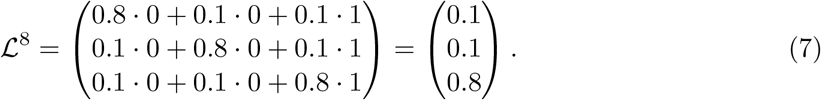

Host 7 is another leaf with deme 2:

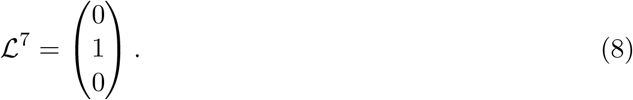

Host 6 is an unsampled individual, whose only offspring is Host 9:

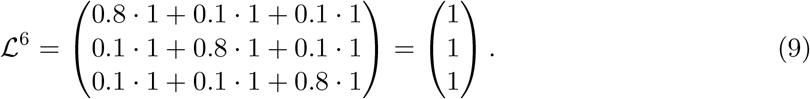

Host 5 is a leaf with deme 1:

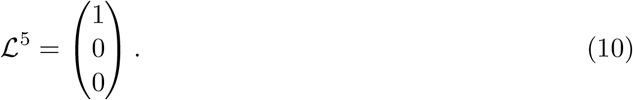

Host 4 is a leaf with no deme data:

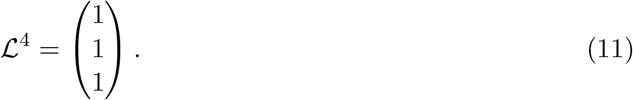

Host 3 has three offspring: Hosts 4, 8, and 7. Additionally, Host 3 is in deme 2, and so we only calculate the second element of the vector:

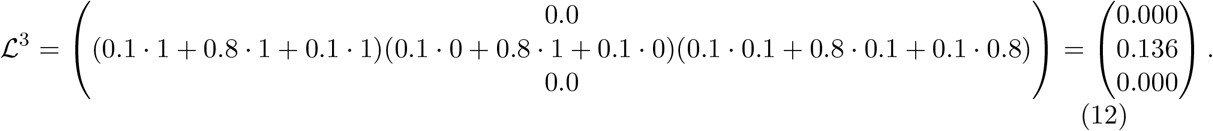

Host 2 has two offspring: Hosts 5 and 6. Host 2 is sampled, but has no deme data, and so all elements of the vector are evaluated:

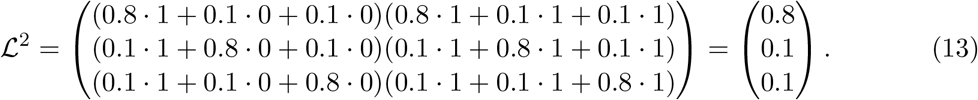

Finally, Host 1 has two offspring: Hosts 2 and 3:

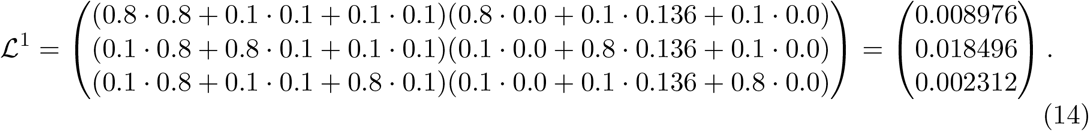

Figure S4B shows the transmission tree annotated with the conditional likelihoods calculated in the dynamic programming algorithm. If we assume that the prior for the deme of the root host is 1*/*3 for the three demes, then the likelihood of the deme is the mean of the values for Host 1. In this case ℒ= 0.009928. This is verified by brute force by calculating

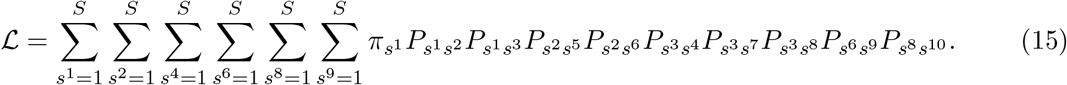

Note that leaves with no deme data do not ultimately contribute to the likelihood, and can therefore be excluded.

### Case where the demes may have different offspring distributions and sampling probabilities

A useful extension would be to allow *R* and/or *π* to change based on deme. For now, let us assume that there are *S* = 2 demes with offspring distribution *α*_1_(*k*) and *α*_2_(*k*). Host infected at time *t* are observed with probability *π*_1_ and *π*_2_ (leading to time-dependent probabilities *ζ*_1_(*t*) and *ζ*_2_(*t*)). Unlike the previous case, the transmission tree likelihood and deme likelihood can not be calculated separately. To evaluate the combined likelihood, we start by calculating the exclusion probabilities as follows.

Define *ω*_1_(*t*) as the exclusion probability of a host infected at time *t* in deme 1, and *ω*_2_(*t*) as the exclusion probability of a host infected at time *t* in deme 2. Assuming that *T* is the cut-off time for observations *ω*_1_(*t*) = *ω*_2_(*t*) = 1 for *t* ≥ *T*. We can then define the following recursive relationships.

The exclusion probability of an offspring from a host in deme 1 infected at time *t* is

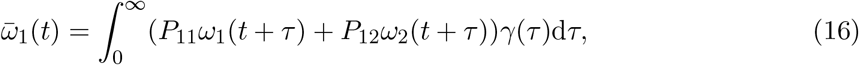

and for deme 2,

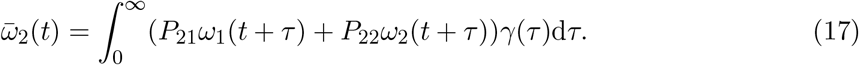

The probability that all offspring from an individual in deme *i* infected at time *t* are excluded is

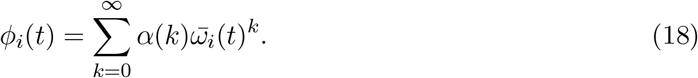

The exclusion probability of an individual in deme *i* infected at time *t* is then

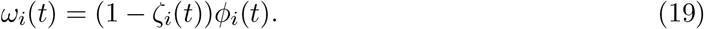

That is, the probability of the host being unobserved and having no included offspring.

As established in Carson et al. (2024), the transmission tree likelihood contribution from an unsampled Host *n* is

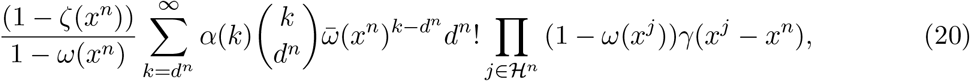

where *x*^*n*^ is the host’s infection time, and *d*^*n*^ is the number of included offspring. If Host *n* is sampled, the likelihood contribution is

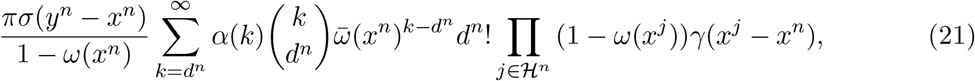

where *y*^*n*^ is the host’s primary observation time, and *σ*(*τ*) is the observation time distribution. Here, we define

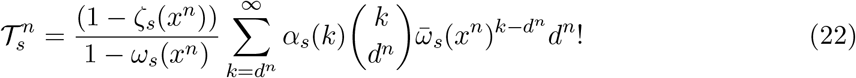

for an unobserved Host *n* in deme *s*, and

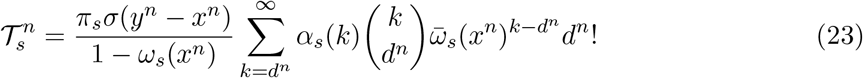

for an observed Host *n* in deme *s*. In addition we define

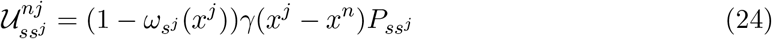

for *j* ∈ ℋ_*n*_ being the offspring of Host *n*, and *s*^*j*^ being the deme of the offspring. Finally, define

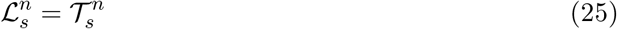

for leaf hosts, and

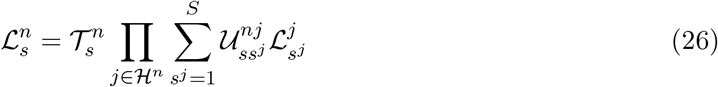

for hosts with offspring. The combined transmission tree and deme likelihood is then calculated using dynamic programming with this replacement definition of 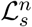.

### Illustrative example

We return to the transmission tree presented in Figure S4A. We include observation times as follows:

| Host | Deme | Infection time | Observation time | Offspring |
| --- | --- | --- | --- | --- |
| 1 | - | 0.0 | - | 2, 3 |
| 2 | - | 2.6 | 3.5 | 5, 6 |
| 3 | 2 | 3.2 | 5.3 | 4, 7, 8 |
| 4 | - | 5.1 | 6.9 | - |
| 5 | 1 | 5.2 | 8.5 | - |
| 6 | - | 5.5 | - | 9 |
| 7 | 2 | 6.3 | 7.1 | - |
| 8 | - | 7.1 | - | 10 |
| 9 | - | 8.9 | 11.0 | - |
| 10 | 3 | 9.8 | 11.6 | - |

We assume that the demes have basic reproduction number *R* equal to 2, 1.5 and 1, respectively, and sampling proportion *π* equal to 0.5, 0.7 and 0.9, respectively. We again set *P* as in Equation (4). Both the generation time distribution and observation time distribution are Gamma distributed with shape 2 and scale 1. The resulting exclusion probabilities are shown in Figure S5.

The resulting conditional likelihoods are as follows:

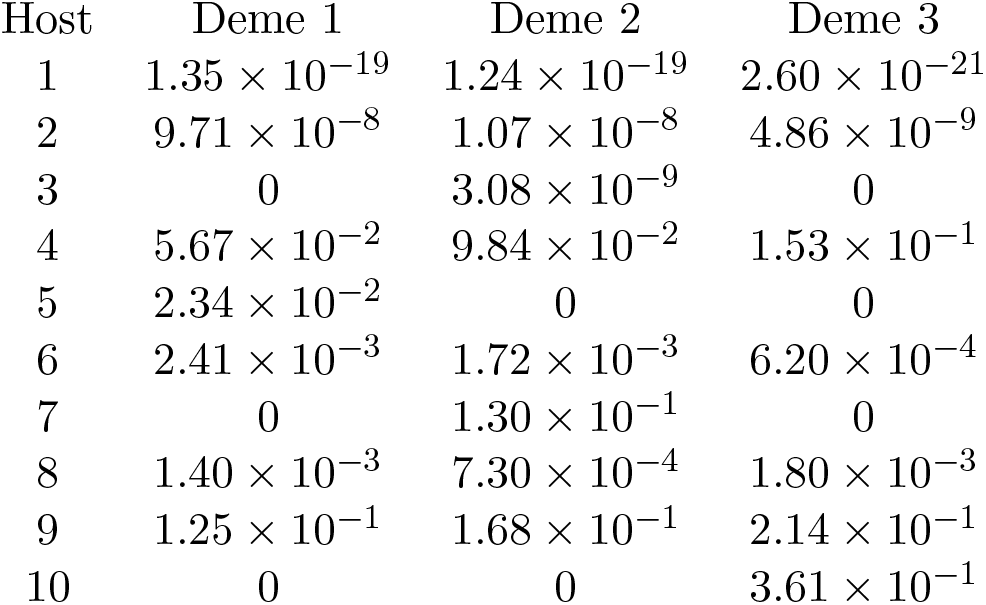

| Host | Deme 1 | Deme 2 | Deme 3 |
| --- | --- | --- | --- |
| 1 | $1.35 \times 10^{-19}$ | $1.24 \times 10^{-19}$ | $2.60 \times 10^{-21}$ |
| 2 | $9.71 \times 10^{-8}$ | $1.07 \times 10^{-8}$ | $4.86 \times 10^{-9}$ |
| 3 | 0 | $3.08 \times 10^{-9}$ | 0 |
| 4 | $5.67 \times 10^{-2}$ | $9.84 \times 10^{-2}$ | $1.53 \times 10^{-1}$ |
| 5 | $2.34 \times 10^{-2}$ | 0 | 0 |
| 6 | $2.41 \times 10^{-3}$ | $1.72 \times 10^{-3}$ | $6.20 \times 10^{-4}$ |
| 7 | 0 | $1.30 \times 10^{-1}$ | 0 |
| 8 | $1.40 \times 10^{-3}$ | $7.30 \times 10^{-4}$ | $1.80 \times 10^{-3}$ |
| 9 | $1.25 \times 10^{-1}$ | $1.68 \times 10^{-1}$ | $2.14 \times 10^{-1}$ |
| 10 | 0 | 0 | $3.61 \times 10^{-1}$ |

The overall likelihood is ℒ = 8.69 *×* 10^−20^, which again is confirmed by using a brute force calculation.

As a further check we recalculate the likelihood under *R* = (2, 2, 2) and *π* = (0.8, 0.8, 0.8). As the demes now have the same offspring distribution and sampling probabilities, we should obtain the same likelihood by taking the product of the transmission tree and deme likelihoods, as in the case where all demes have the same offspring distribution and sampling probabilities. We find that both approaches do indeed return the same likelihood.

### Implementation

We implemented the methods above into a new R package called TransPhylo2, which extends TransPhyloMulti (Carson et al., 2024) and therefore inherits the same advantages over the previous implementation of TransPhylo (Didelot et al., 2017) in terms of allowing multiple samples per host and relaxing the assumption of a complete transmission bottleneck. TransPhylo2 is available at https://github.com/DrJCarson/TransPhylo2. This repository also contains all the code and data needed to reproduce all results shown in this paper. The R package ape was used to store, manipulate and visualise phylogenetic trees (Paradis and Schliep, 2019).

## Data Availability

All data produced are available online at https://github.com/DrJCarson/TransPhylo2

## ACKNOWLEDGEMENTS

We acknowledge funding from the National Institute for Health Research (NIHR) Health Protection Research Unit in Genomics and Enabling Data (grant number NIHR200892).

## References

Baele G, Suchard MA, Rambaut A, Lemey P. 2017. Emerging concepts of data integration in pathogen phylodynamics. Systematic biology. 66:e47–e65.

Bataille A, Van Der Meer F, Stegeman A, Koch G. 2011. Evolutionary Analysis of Inter-Farm Transmission Dynamics in a Highly Pathogenic Avian Influenza Epidemic. PLoS Pathogens. 7:e1002094.

Biek R, Pybus OG, Lloyd-Smith JO, Didelot X. 2015. Measurably evolving pathogens in the genomic era. Trends in Ecology & Evolution. 30:306–313.

Bouckaert R, Vaughan TG, Fourment M, Gavryushkina A, Heled J, Denise K, Maio ND, Matschiner M, Ogilvie H, Plessis L, et al. (11 co-authors). 2019. BEAST 2.5 : An Advanced Software Platform for Bayesian Evolutionary Analysis. PLoS computational biology. 15:e1006650.

Brooks SPB, Gelman AG. 1998. General methods for monitoring convergence of iterative simulations. Journal of computational and graphical statistics. 7:434–455.

Brooks-Pollock E, Danon L, Jombart T, Pellis L. 2021. Modelling that shaped the early COVID-19 pandemic response in the UK. Philosophical Transactions of the Royal Society B: Biological Sciences. 376:20210001.

Bruchfeld J, Correia-Neves M, Källenius G. 2015. Tuberculosis and HIV Coinfection. Cold Spring Harbor Perspectives in Medicine. 5:a017871.

Campbell F, Strang C, Ferguson N, Cori A, Jombart T. 2018. When are pathogen genome sequences informative of transmission events? PLOS Pathogens. 14:e1006885.

Carson J, Keeling M, Wyllie D, Ribeca P, Didelot X. 2024. Inference of infectious disease transmission through a relaxed bottleneck using multiple genomes per host. Molecular Biology and Evolution. 41:msad288.

Chan CH, McCabe CJ, Fisman DN. 2012. Core groups, antimicrobial resistance and rebound in gonorrhoea in North America. Sexually Transmitted Infections. 88:200–204.

Chis Ster I, Singh BK, Ferguson NM. 2009. Epidemiological inference for partially observed epidemics: The example of the 2001 foot and mouth epidemic in Great Britain. Epidemics. 1:21–34.

Chitwood MH, Corbett EL, Ndhlovu V, Sobkowiak B, Colijn C, Andrews JR, Burke RM, Cudahy PG, Dodd PJ, Imai-Eaton JW, et al. (23 co-authors). 2024. Distribution and transmission of M. tuberculosis in a high-HIV prevalence city in Malawi: A genomic and spatial analysis. medRxiv. medRxiv:2024.05.17.24307525.

Cori A, Ferguson NM, Fraser C, Cauchemez S. 2013. A new framework and software to estimate time-varying reproduction numbers during epidemics. American journal of epidemiology. 178:1505–12.

Craig AP, Gray RT, Edwards JL, Apicella MA, Jennings MP, Wilson DP, Seib KL. 2015. The potential impact of vaccination on the prevalence of gonorrhea. Vaccine. 33:4520–4525.

Croucher NJ, Didelot X. 2015. The application of genomics to tracing bacterial pathogen transmission. Current Opinion in Microbiology. 23:62–67.

De Maio N, Wu CH, O’Reilly KM, Wilson D. 2015. New Routes to Phylogeography: A Bayesian Structured Coalescent Approximation. PLoS Genetics. 11:e1005421.

Didelot X, Fraser C, Gardy J, Colijn C. 2017. Genomic infectious disease epidemiology in partially sampled and ongoing outbreaks. Molecular Biology and Evolution. 34:997–1007.

Didelot X, Gardy J, Colijn C. 2014. Bayesian inference of infectious disease transmission from whole genome sequence data. Molecular Biology and Evolution. 31:1869–1879.

Didelot X, Kendall M, Xu Y, White PJ, McCarthy N. 2021. Genomic Epidemiology Analysis of Infectious Disease Outbreaks Using TransPhylo. Current Protocols. 1:e60.

Didelot X, Parkhill J. 2022. A scalable analytical approach from bacterial genomes to epidemiology. Philosophical Transactions of the Royal Society B: Biological Sciences. 377:20210246.

Drummond AJ, Suchard MA, Xie D, Rambaut A. 2012. Bayesian phylogenetics with BEAUti and the BEAST 1.7. Molecular Biology and Evolution. 29:1969–1973.

Duault H, Durand B, Canini L. 2022. Methods Combining Genomic and Epidemiological Data in the Reconstruction of Transmission Trees: A Systematic Review. Pathogens. 11:252.

Eldholm V, Monteserin J, Rieux A, Lopez B, Sobkowiak B, Ritacco V, Balloux F. 2015. Four decades of transmission of a multidrug-resistant Mycobacterium tuberculosis outbreak strain. Nature communications. 6:7119.

Eldholm V, Rieux A, Monteserin J, Lopez JM, Palmero D, Lopez B, Ritacco V, Didelot X, Balloux F. 2016. Impact of HIV co-infection on the evolution and transmission of multidrug-resistant tuberculosis. eLife. 5:e16644.

Farrington CP, Kanaan MN, Gay NJ. 2003. Branching process models for surveillance of infectious diseases controlled by mass vaccination. Biostatistics (Oxford, England). 4:279–95.

Felsenstein J. 1973. Maximum Likelihood and Minimum-Steps Methods for Estimating Evolutionary Trees from Data on Discrete Characters. Systematic Zoology. 22:240–49.

Felsenstein J. 1981. Evolutionary trees from DNA sequences: A maximum likelihood approach. Journal of Molecular Evolution. 17:368–376.

Fingerhuth SM, Bonhoėer S, Low N, Althaus CL. 2016. Antibiotic-Resistant Neisseria gonorrhoeae Spread Faster with More Treatment, Not More Sexual Partners. PLOS Pathogens. 12:e1005611.

Fraser C, Riley S, Anderson RM, Ferguson NM. 2004. Factors that make an infectious disease outbreak controllable. Proceedings of the National Academy of Sciences. 101:6146–6151.

Glatt-Holtz NE, Holbrook AJ, Krometis JA, Mondaini CF. 2024. Parallel MCMC algorithms: Theoretical foundations, algorithm design, case studies. Transactions of Mathematics and Its Applications. 8:tnae004.

Grassly NC, Fraser C. 2008. Mathematical models of infectious disease transmission. Nature Reviews Microbiology. 6:477–87.

Green PJ. 1995. Reversible Jump Markov Chain Monte Carlo Computation and Bayesian Model Determination. Biometrika. 82:711–732.

Hall M, Woolhouse M, Rambaut A. 2015. Epidemic Reconstruction in a Phylogenetics Framework: Transmission Trees as Partitions of the Node Set. PLOS Computational Biology. 11:e1004613.

Hatherell HA, Didelot X, Pollock SL, Tang P, Crisan A, Johnston JC, Colijn C, Gardy J. 2016. Declaring a tuberculosis outbreak over with genomic epidemiology. Microbial Genomics. 1:10.1099/mgen.0.000060.

Hollingsworth TD. 2009. Controlling infectious disease outbreaks: Lessons from mathematical modelling. Journal of Public Health Policy. 30:328–341.

Jewell CP, Keeling MJ, Roberts GO. 2009. Predicting undetected infections during the 2007 foot-and-mouth disease outbreak. Journal of The Royal Society Interface. 6:1145–1151.

Jombart T, Cori A, Didelot X, Cauchemez S, Fraser C, Ferguson N. 2014. Bayesian Reconstruction of Disease Outbreaks by Combining Epidemiologic and Genomic Data. PLoS Computational Biology. 10:e1003457.

Klinkenberg D, Backer JA, Didelot X, Colijn C, Wallinga J. 2017. Simultaneous inference of phylogenetic and transmission trees in infectious disease outbreaks. PLoS Computational Biology. 13:e1005495.

Layan M, Müller NF, Dellicour S, De Maio N, Bourhy H, Cauchemez S, Baele G. 2023. Impact and mitigation of sampling bias to determine viral spread: Evaluating discrete phylogeography through CTMC modeling and structured coalescent model approximations. Virus Evolution. 9:vead010.

Lemey P, Rambaut A, Drummond AJ, Suchard MA. 2009. Bayesian phylogeography finds its roots. PLoS computational biology. 5:e1000520.

Magnani R, Sabin K, Saidel T, Heckathorn D. 2005. Review of sampling hard-to-reach and hidden populations for HIV surveillance. AIDS. 19:S67–S72.

Mollentze N, Nel LH, Townsend S, le Roux K, Hampson K, Haydon DT, Soubeyrand S. 2014. A Bayesian approach for inferring the dynamics of partially observed endemic infectious diseases from space-time-genetic data. Proceedings of the Royal Society B: Biological Sciences. 281:20133251.

Morelli MJ, Thébaud G, Chadœuf J, King DP, Haydon DT, Soubeyrand S. 2012. A Bayesian Inference Framework to Reconstruct Transmission Trees Using Epidemiological and Genetic Data. PLoS Computational Biology. 8:e1002768.

O’Neill PD. 2002. A tutorial introduction to Bayesian inference for stochastic epidemic models using Markov chain Monte Carlo methods. Mathematical biosciences. 180:103–14.

O’Neill PD, Roberts GO. 1999. Bayesian inference for partially observed stochastic epidemics. Journal of the Royal Statistical Society: Series A. 162:121–129.

Paradis E, Schliep K. 2019. Ape 5.0: An environment for modern phylogenetics and evolutionary analyses in R. Bioinformatics. 35:526–528.

Rieux A, Balloux F. 2016. Inferences from tip-calibrated phylogenies: A review and a practical guide. Molecular Ecology. 25:1911–1924.

Schwedes T, Calderhead B. 2021. Rao-Blackwellised parallel MCMC. Aistats. 130.

Séraphin MN, Didelot X, Nolan DJ, May JR, Khan MSR, Murray ER, Salemi M, Morris JG, Lauzardo M. 2018. Genomic Investigation of a Mycobacterium tuberculosis Outbreak Involving Prison and Community Cases in Florida, United States. American Journal of Tropical Medicine and Hygiene. 99:867–874.

Shu Y, McCauley J. 2017. GISAID: Global initiative on sharing all influenza data – from vision to reality. Eurosurveillance. 22.

Sisson SA. 2005. Transdimensional Markov chains: A decade of progress and future perspectives. Journal of the American Statistical Association. 100:1077–1089.

Sobkowiak B, Romanowski K, Sekirov I, Gardy JL, Johnston JC. 2023. Comparing Mycobacterium tuberculosis transmission reconstruction models from whole genome sequence data. Epidemiology and Infection. 151:e105.

Stegeman A, Bouma A, Elbers ARW, de Jong MCM, Nodelijk G, de Klerk F, Koch G, van Boven M. 2004. Avian Influenza A Virus (H7N7) Epidemic in The Netherlands in 2003: Course of the Epidemic and E?ectiveness of Control Measures. The Journal of Infectious Diseases. 190:2088–2095.

Syed S, Bouchard-Côté A, Deligiannidis G, Doucet A. 2022. Non-Reversible Parallel Tempering: A Scalable Highly Parallel MCMC Scheme. Journal of the Royal Statistical Society Series B: Statistical Methodology. 84:321–350.

van Dyk DA, Meng XL. 2001. The Art of Data Augmentation. Journal of Computational and Graphical Statistics. 10:1–50.

Wallinga J, Teunis P. 2004. Di?erent Epidemic Curves for Severe Acute Respiratory Syndrome Reveal Similar Impacts of Control Measures. American Journal of Epidemiology. 160:509–516.

Whittles LK, Didelot X, White PJ. 2022. Public health impact and cost-e?ectiveness of gonorrhoea vaccination: An integrated transmission-dynamic health-economic modelling analysis. Lancet Infectious Diseases. 22:1030–1041.

Whittles LK, White PJ, Didelot X. 2019. A dynamic power-law sexual network model of gonorrhoea outbreaks. PLoS Computational Biology. 15:e1006748.

Whittles LK, White PJ, Didelot X. 2020. Assessment of the Potential of Vaccination to Combat Antibiotic Resistance in Gonorrhea: A Modeling Analysis to Determine Preferred Product Characteristics. Clinical Infectious Diseases. 71:1912–1919.

Ypma RJF, Bataille AMA, Stegeman A, Koch G, Wallinga J, van Ballegooijen WM. 2012. Unravelling transmission trees of infectious diseases by combining genetic and epidemiological data. Proceedings of the Royal Society B. 279:444–450.

Ypma RJF, van Ballegooijen WM, Wallinga J. 2013. Relating Phylogenetic Trees to Transmission Trees of Infectious Disease Outbreaks. Genetics. 195:1055–1062.

