## Supplementary Material for "Incorporating epidemiological data into the genomic analysis of partially sampled infectious disease outbreaks"

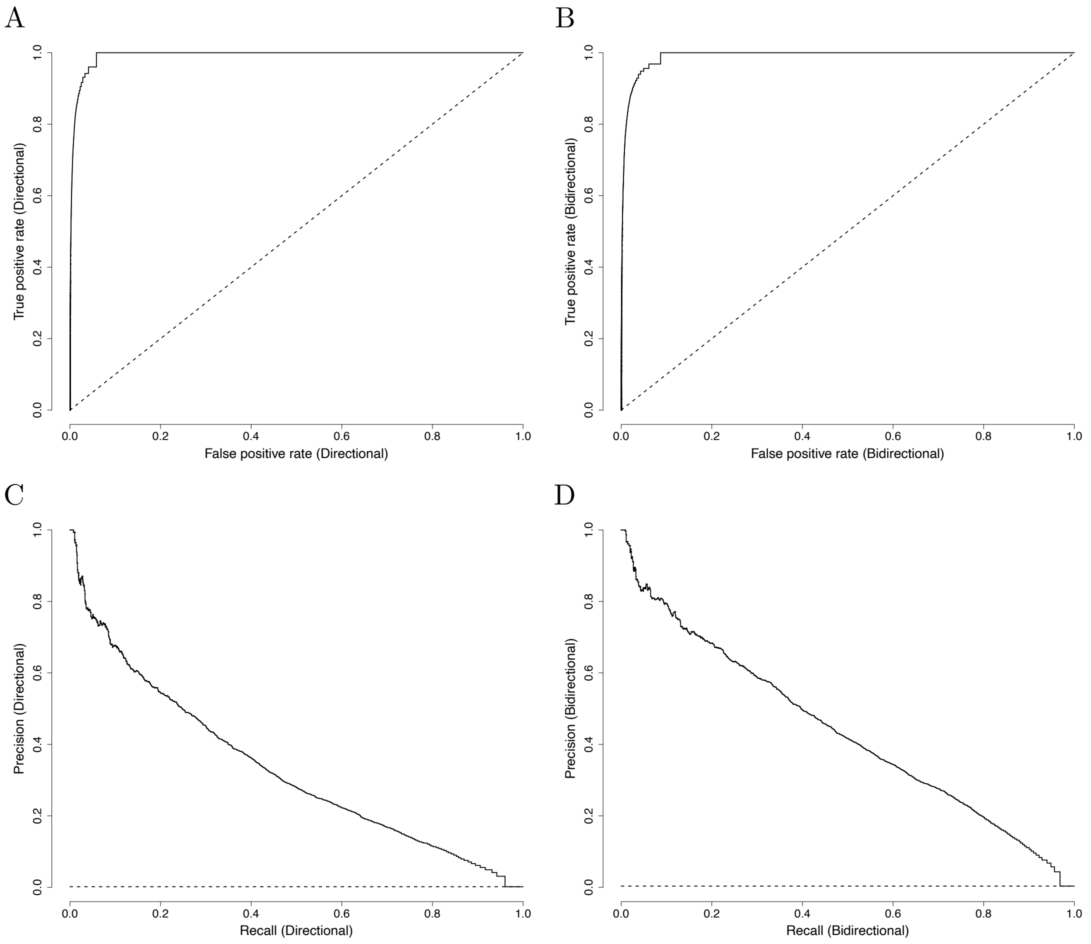


Figure S1: Receiver Operating Characteristic (ROC) curves for directional (A) and bidirectional (B) transmission links averaged across the 50 benchmarking simulations. Precision-Recall (PR) curves for directional (C) and bidirectional (D) transmission links averaged across the 50 benchmarking simulations.

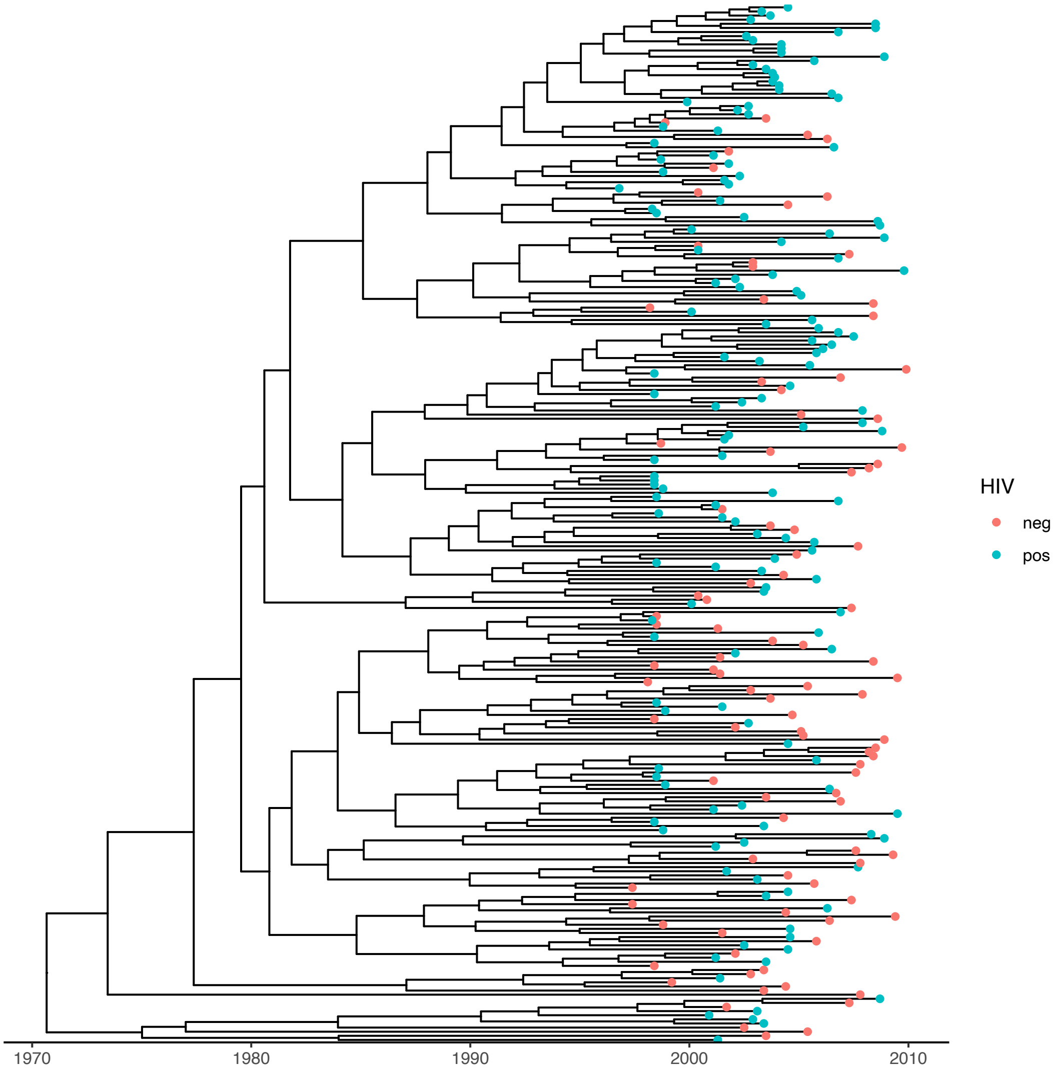


Figure S2: Dated tree used in the tuberculosis application, with leaves colored by HIV status.

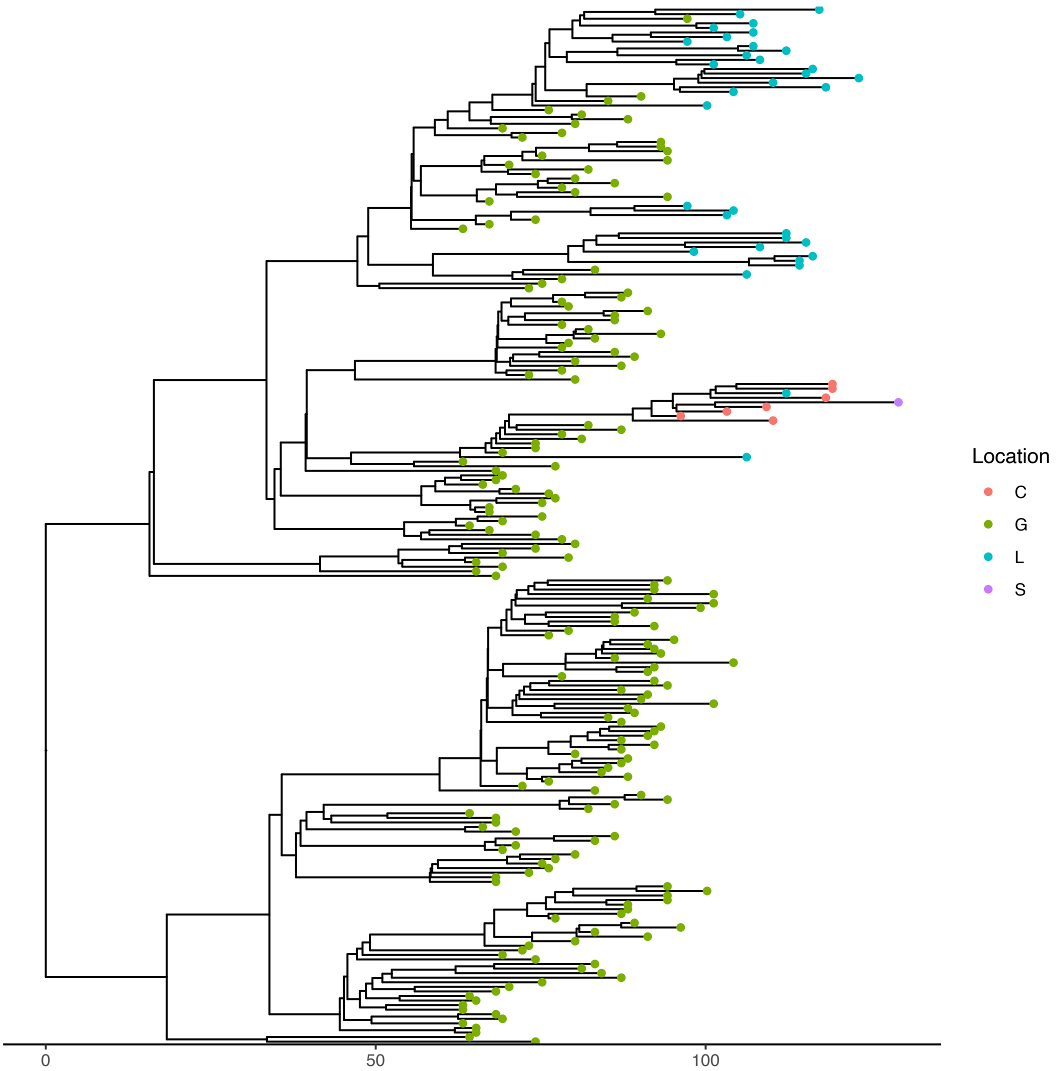


Figure S3: Dated tree used in the H7N7 application, with leaves colored by location.

A

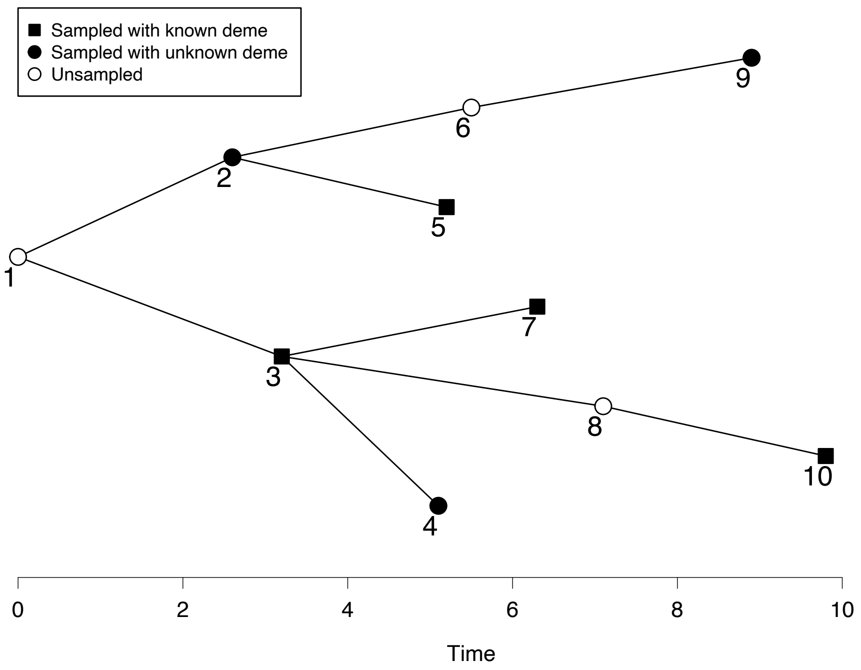


B

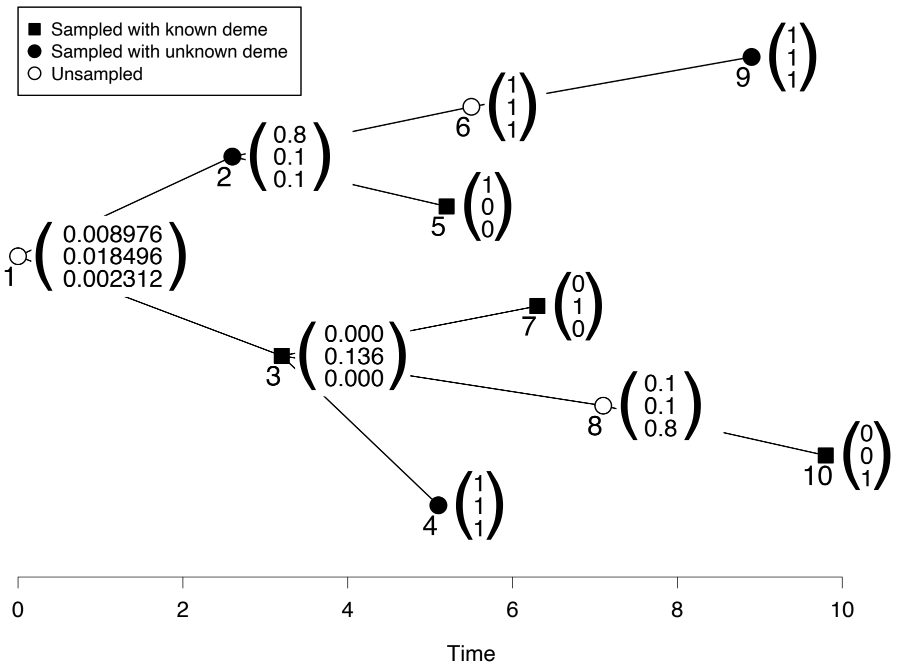


Figure S4: (A) Example transmission tree with 10 hosts. Three hosts are unsampled, three hosts are sampled without any deme data, and four hosts are sampled with deme data. (B) Conditional likelihoods in the dynamic programming algorithm with three demes, shown as vectors by each host representing the conditional likelihood at the three demes. The final likelihood for the demes is the mean of the three values for Host 1.

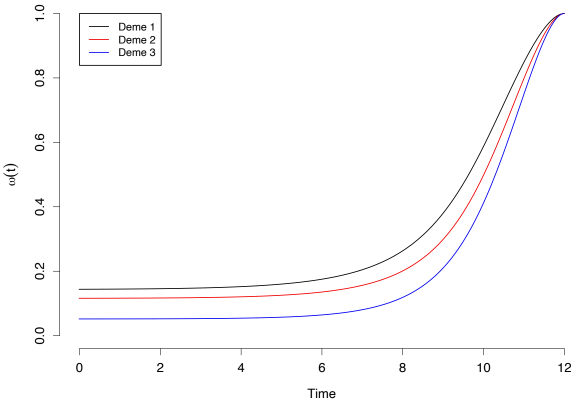


Figure S5: Exclusion probabilities through time for the illustrative example.
